# Composite Artificial Intelligence–Enabled Electrocardiogram for Detection and Prediction of Structural Heart Disease

**DOI:** 10.64898/2026.07.21.26358539

**Authors:** Hak Seung Lee, Sora Kang, Min Sung Lee, Ambarish Pandey, Minkwan Kim, Jong-Hwan Jang, Yong-Yeon Jo, Jaehyun Lim, Jeong Min Son, Kyung Su Kim, Joon-myoung Kwon, Seung-Pyo Lee, Kyung-Hee Kim

## Abstract

**Background:** Structural heart disease (SHD) drives heart failure and cardiovascular mortality but remains underdiagnosed, and echocardiography is limited as a population-level screening tool.

**Objectives:** We evaluated whether a composite artificial intelligence–enabled electrocardiogram (AI-ECG), combining independently developed models for left ventricular systolic (LVSD) and diastolic dysfunction (LVDD), identifies prevalent and predicts incident SHD across diverse populations.

**Methods:** In this multinational cohort study, detection was assessed cross-sectionally in a Korean clinical cohort (Incheon Sejong Hospital) and a US dataset (Columbia University Irving Medical Center), and incident risk was assessed in the Korean cohort and the UK Biobank among individuals without baseline SHD or heart failure. Adults with paired ECG and echocardiography were analyzed for detection, with the composite defined as positive on either model. SHD comprised reduced left ventricular ejection fraction, moderate or severe valvular disease, left ventricular hypertrophy, or pulmonary hypertension. Detection was assessed by sensitivity and specificity, and incident risk by Cox models and the C statistic.

**Results:** Among 46,082 and 36,286 participants in the two detection cohorts, the composite detected SHD with sensitivity of 71.8% and 76.1% and specificity of 88.3% and 70.1%, with positivity across all phenotypes. Among at-risk individuals, composite positivity was associated with incident SHD (hazard ratios, 3.75 and 2.75), with C statistics of 0.69 to 0.78.

**Conclusions:** A composite AI-ECG identified prevalent and predicted incident SHD across multinational cohorts, capturing signals beyond its training targets and supporting its potential as a scalable cardiovascular screening tool; whether ECG-based risk stratification improves outcomes requires prospective evaluation.

## Introduction

Structural heart disease (SHD) — including valvular disease, ventricular dysfunction, ventricular hypertrophy, and pulmonary hypertension — is a major contributor to heart failure (HF) and premature mortality, and its burden continues to grow with population aging.(1–5)

Yet SHD remains substantially underdiagnosed, with many patients identified only after disease progression. Echocardiography is required for definitive diagnosis, but its cost, limited accessibility, and need for specialized expertise constrain its use as a population-level screening tool, leaving a critical need for scalable early identification and risk stratification — particularly as SHD develops along a continuum from subclinical alterations to overt disease.(6)

Artificial intelligence–enabled electrocardiogram (AI-ECG) is increasingly recognized as a digital biomarker reflecting structural and functional myocardial abnormalities.(7) Beyond detecting individual conditions such as reduced ejection fraction, hypertrophy, and valvular disease, composite models — including rECHOmmend, EchoNext, and PRESENT-SHD — reframe SHD as a clinically meaningful risk state warranting echocardiographic referral.(8–10) These tools nonetheless remain anchored to condition-specific labels. Emerging evidence, however, indicates that AI-ECG models capture signals broader than their nominal targets: models trained for distinct conditions show overlapping phenotype associations and detect off-target disease, and a binary composite of AI-ECG models for left ventricular systolic (LVSD) and diastolic dysfunction (LVDD) independently predicted incident HF across US population-based cohorts.(11,12)

Whether this paradigm generalizes to prevalent and incident SHD across diverse populations remains unclear, as existing evidence is largely confined to single-cohort or US-based populations, a single model family, and HF as the outcome. We therefore evaluated whether a composite AI-ECG combining independent, clinically deployed LVSD and LVDD models identifies prevalent SHD and predicts incident SHD across three multinational cohorts spanning Korea, the United States, and the United Kingdom.

## Methods

### Study Design and Data Sources

This study evaluated a composite AI-ECG across three complementary multinational cohorts: Incheon Sejong Hospital (ISH; Republic of Korea), Columbia University Irving Medical Center (CUIMC; United States), and the UK Biobank (UKB; United Kingdom). The ISH and CUIMC cohorts were used for SHD detection, and the ISH and UKB cohorts for new-onset SHD. The overall design, cohort-specific data curation, and analytic workflow are summarized in Figure 1; full curation and ECG-acquisition details are provided in the Supplementary Methods.

**Figure 1.**
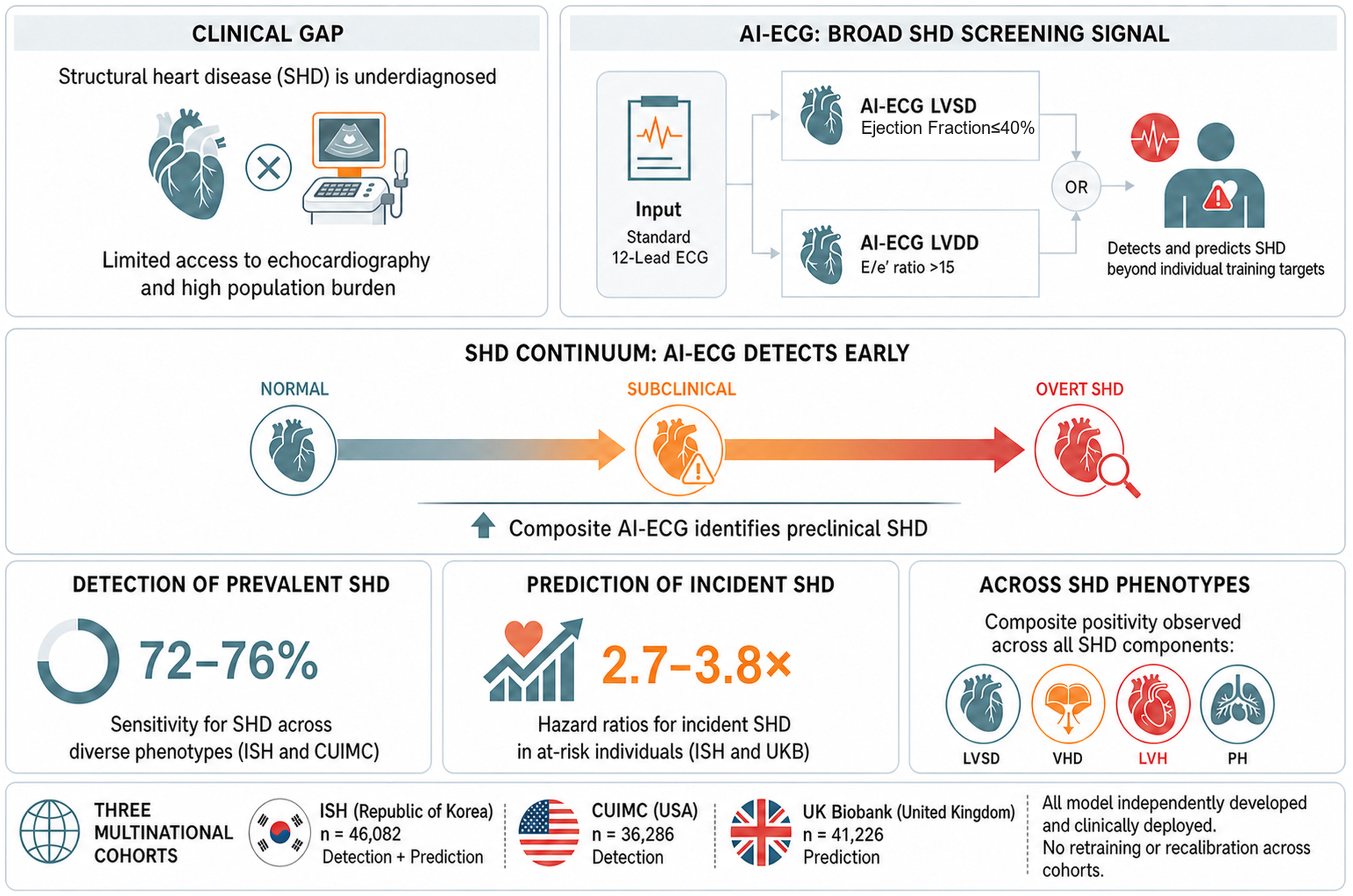
Study cohorts, data curation, and analytic workflow Three independent cohorts were included in this study: ISH, CUIMC, and the UKB. Each cohort underwent cohort-specific inclusion and exclusion procedures to generate curated analytic datasets. The curated datasets were subsequently used for two distinct analytic objectives. For cross-sectional detection of SHD, ECG–echocardiography–paired data from the ISH and CUIMC curated datasets were analyzed. For longitudinal prediction of new-onset SHD, at-risk participants without baseline SHD or heart failure from the ISH and UKB curated datasets were followed over time. ISH contributed to both analyses through separate cross-sectional and longitudinal datasets, whereas CUIMC and UKB were used exclusively for SHD detection and new-onset SHD prediction, respectively. ISH, Incheon Sejong Hospital; CUIMC, Columbia University Irving Medical Center; UKB, UK Biobank; SHD, structural heart disease; ECG, electrocardiogram.

The ISH cohort comprised adults (≥18 years) who underwent 12-lead ECG and transthoracic echocardiography within 14 days between January 2016 and December 2024, with the most recent eligible pair selected per participant. The CUIMC cohort was a publicly released, de-identified dataset of 100,000 ECGs from 36,286 adults paired with echocardiography-derived labels, used exclusively for detection.(8) The UKB is a prospective, population-based cohort of more than 500,000 adults aged 40–69 years at enrollment (2006–2010), with a subset undergoing 12-lead ECG (2014–2021) and longitudinal linkage to National Health Service and death-registry records; participants without baseline SHD were analyzed for incident SHD (application 89372).(13,14)

The study was approved by the Institutional Review Board of ISH; given the retrospective use of previously collected and publicly available data, the requirement for informed consent was waived.

### Study Outcomes

SHD was defined as the presence of any of reduced left ventricular ejection fraction (LVEF), moderate or severe valvular heart disease (VHD), left ventricular hypertrophy (LVH), or pulmonary hypertension (PH) on echocardiography, based on clinical interpretations performed in accordance with contemporary echocardiographic guidelines; component-specific thresholds are detailed in Supplementary Table 1.(8–10,15–17) Reduced LVEF was defined as an LVEF <50%. Notably, the AI-ECG LVSD model was originally trained to detect an LVEF ≤40%; this mismatch with the SHD outcome definition (<50%) was retained intentionally to test whether the model, when combined with the AI-ECG LVDD model, captures the broader SHD spectrum beyond its original training target.

For the prediction analyses, individuals with no baseline SHD and no prior HF constituted the at-risk population. In ISH, baseline SHD was excluded by review of echocardiography reports, prior HF was identified from the electronic medical record (ICD-10 code I50), and incident SHD was defined as the first echocardiography-confirmed event during follow-up. In UKB, where echocardiography was unavailable, at-risk status and incident events were ascertained from ICD-10 codes in linked national health records, with the systolic-dysfunction component operationalized as incident HF rather than echocardiographic reduced LVEF; component codes are listed in Supplementary Table 1. In both cohorts, only individuals with ≥6 months of follow-up were included, censored at the first occurrence of incident SHD, incident HF, death, or the end of follow-up.

### AI-ECG models

Two AI-ECG models—for LVSD and LVDD—were used, and the composite AI-ECG was defined as a positive result on either. Both were developed by Medical AI Co., Ltd., received regulatory approval from the Korean Ministry of Food and Drug Safety, and were validated in prior studies;(18–22) they use digital signals from standard 12-lead ECGs alone, without additional clinical variables. Both were derived using a two-stage framework based on a previously established ECG foundation model.(23) The LVSD model was trained to identify an LVEF ≤40% and the LVDD model an echocardiographic septal E/e′ ratio >15; each generates a continuous probability score (0–100), with binary thresholds set by the Youden index in the derivation cohort. The models were applied without retraining or recalibration, and their development datasets had no overlap with any cohort in this study, ensuring independent external validation. Foundation-model pretraining and fine-tuning and a separately developed end-to-end SHD benchmark model are detailed in the eMethods in Supplement 1.

### Statistical analysis

Continuous variables are summarized as medians with interquartile ranges and categorical variables as counts and percentages. Detection performance was evaluated using sensitivity, specificity, positive predictive value (PPV), and negative predictive value (NPV); because PPV and NPV are prevalence-dependent, cross-cohort comparisons were interpreted in the context of differing baseline SHD prevalence. Ninety-five percent CIs for all performance metrics were estimated from 1000 bootstrap resamples. Subgroup analyses were performed across strata defined by age, sex, medical history, and ECG rhythm (eMethods in Supplement 1).

For the prediction analyses, the earliest available ECG served as the baseline; the at-risk population therefore included both true-negative and false-positive baseline composite results. The association between baseline composite positivity and incident SHD was evaluated using Cox proportional hazards models adjusted for age and sex, with hazard ratios (HRs) reported, and discrimination was assessed using the Harrell C statistic. The two longitudinal cohorts were analyzed in parallel rather than pooled—ISH providing imaging-confirmed ascertainment in a clinical population and UKB administrative-code–based ascertainment in a population-based cohort—to assess reproducibility across complementary outcome definitions. In an exploratory analysis, the joint distribution of LVSD and LVDD scores was visualized as two-dimensional heatmaps of SHD-phenotype prevalence.

All tests were 2-sided, with significance set at P < .05. Analyses used Python 3.11.2 and R 4.2.0. The study followed the TRIPOD+AI (Transparent Reporting of a multivariable prediction model for Individual Prognosis Or Diagnosis + Artificial Intelligence) reporting guideline (Supplementary Table 2).

## Results

### Study Population

The detection analyses included 46,082 participants from ISH and 36,286 from CUIMC. Baseline characteristics are summarized in Supplementary Table 3. The median age was 60.0 years (IQR, 50.0–71.0) in ISH and 64.0 years (IQR, 52.0–75.0) in CUIMC, and women comprised 48.0% and 50.5%, respectively. The ISH cohort was ethnically homogeneous (all Asian), whereas CUIMC was racially and ethnically diverse; baseline comorbidity data were available only for ISH (hypertension, 29.2%; diabetes, 8.5%; prior myocardial infarction, 3.2%; prior HF, 18.0%; atrial fibrillation, 10.5%).

Echocardiography showed a higher SHD burden in CUIMC than ISH. Mean LVEF was 62.6% (SD, 9.1) in ISH and 53.4% (SD, 13.6) in CUIMC, and composite SHD prevalence was 12.7% vs 43.6%. Component prevalences for reduced LVEF, moderate or severe VHD, LVH, and PH were 7.5%, 3.7%, 1.7%, and 2.9% in ISH and 20.6%, 15.9%, 18.8%, and 13.4% in

CUIMC, respectively.

### Detection of SHD

Detection performance of the composite AI-ECG and the individual LVSD and LVDD models, assessed against echocardiographic diagnoses as the reference standard, is summarized in Table 1. At the predefined thresholds, 19.3% of participants in ISH and 50.1% in CUIMC screened positive. In ISH, the composite achieved a sensitivity of 71.8% (95% CI, 70.6–72.9) and specificity of 88.3% (95% CI, 88.0–88.6), with a PPV of 47.2% (95% CI, 46.2–48.3) and NPV of 95.6% (95% CI, 95.4–95.8). In CUIMC, sensitivity was 76.1% (95% CI, 75.4–76.7) and specificity 70.1% (95% CI, 69.5–70.8), with a PPV of 66.3% (95% CI, 65.6–67.0) and NPV of 79.1% (95% CI, 78.6–79.7); the lower PPV and higher NPV in ISH were consistent with its lower SHD prevalence (12.7% vs 43.6%). The individual models showed consistent profiles across cohorts: the LVSD model had higher specificity with lower sensitivity, and the LVDD model higher sensitivity with moderate specificity. At matched sensitivity, a separately developed end-to-end AI-ECG SHD model achieved specificity comparable to the composite in both cohorts (Supplementary Table 4).

**Table 1.**
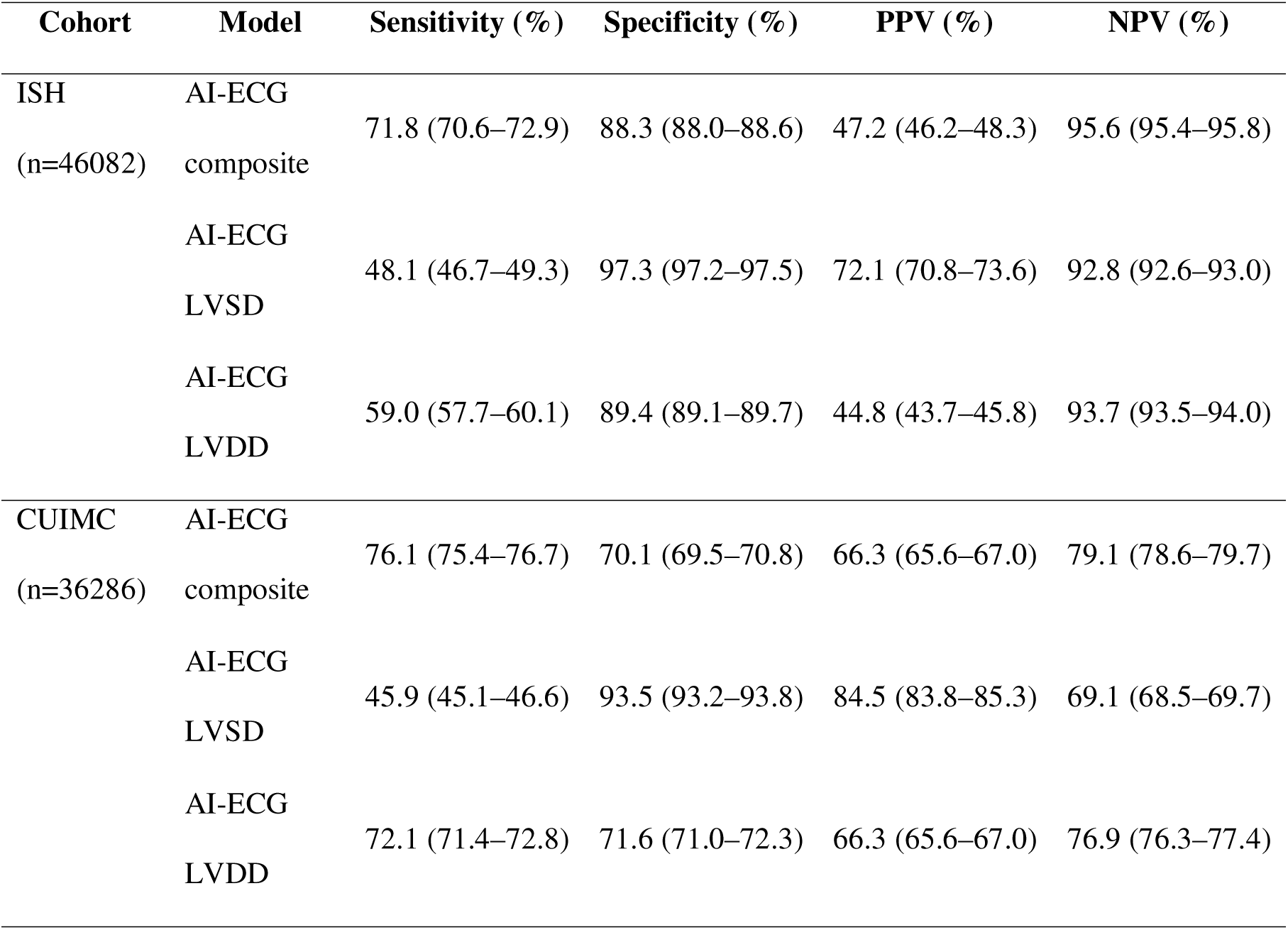
Performance of AI-ECG models for detection of SHD in the ISH and CUIMC cohorts.

The distribution of SHD components differed substantially by composite status in both cohorts (Figure 2). In ISH, most individuals with composite SHD and with each individual phenotype were composite positive, with the highest proportions for reduced LVEF and PH; in CUIMC, composite positivity remained predominant across all phenotypes despite the greater overall burden. Stratification by individual model outputs revealed phenotype-specific patterns (Supplementary Figure 3): LVSD-model signals predominated in the reduced-LVEF phenotype, whereas LVDD-model signals were relatively more prominent in the other phenotypes (VHD, LVH, and PH). The proportion classified composite positive increased stepwise with the number of co-occurring SHD components (Supplementary Figure 4), and AI-ECG–defined strata showed graded differences in echocardiographic systolic function, valvular severity, wall thickness, and pulmonary pressure (Supplementary Figure 5). In exploratory analyses, the joint distribution of LVSD and LVDD scores illustrated distinct phenotype prevalence patterns across the two-dimensional score space (Supplementary Figure 6); relative contributions of each model to composite detection are detailed in Supplementary Table 5 and Supplementary Figure 7.

**Figure 2.**
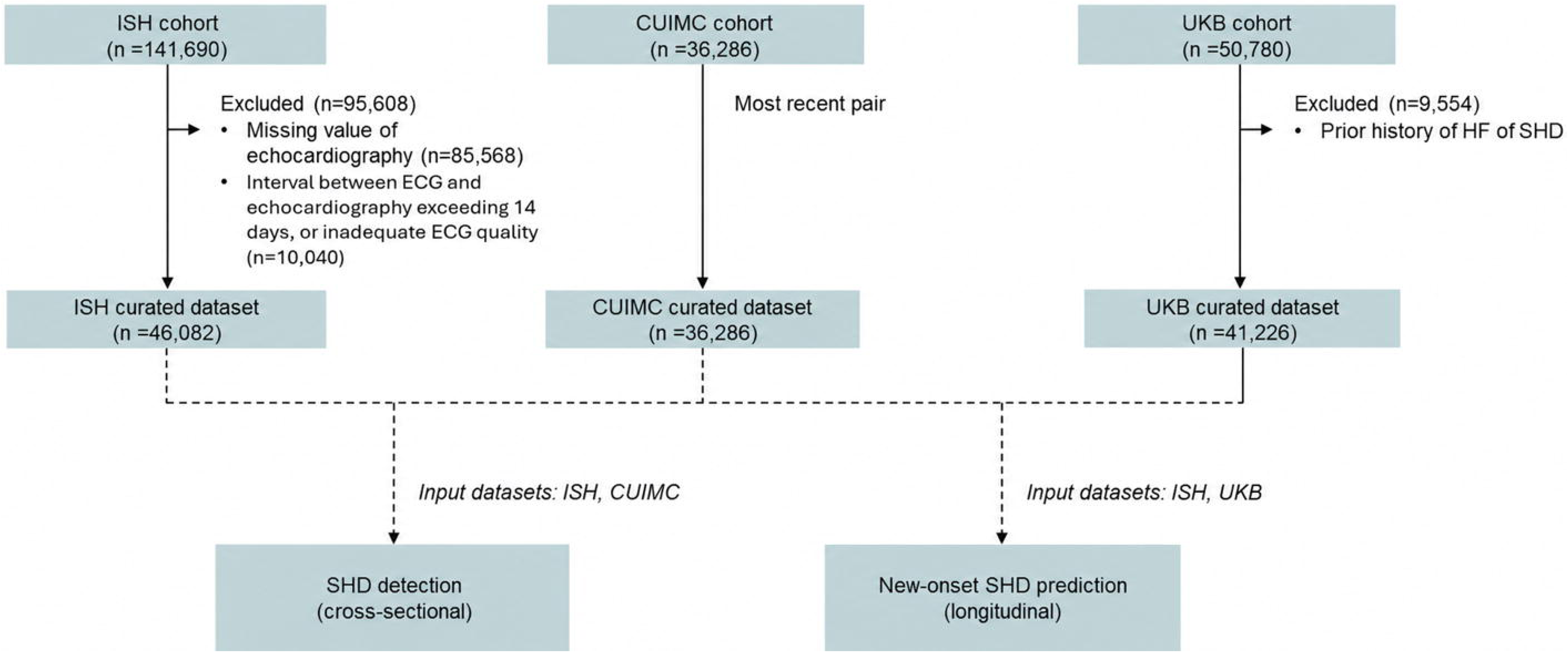
Component-wise prevalence of SHD stratified by AI-ECG composite status Stacked bar charts show the prevalence of composite SHD and its individual components—LVSD, moderate or severe VHD, LVH, and PH—in the ISH and CUIMC cohorts. The height of each bar represents the proportion of the total cohort with the corresponding phenotype. Each bar is partitioned according to AI-ECG composite–positive and –negative status. Percentages displayed within bars indicate the proportion of individuals with each phenotype who were classified as AI-ECG composite positive or negative, whereas percentages displayed above bars indicate the overall prevalence of each phenotype in the total cohort. AI-ECG, artificial intelligence–enabled electrocardiogram; SHD, structural heart disease; LVSD, left ventricular systolic dysfunction; VHD, valvular heart disease; LVH, left ventricular hypertrophy; PH, pulmonary hypertension; ISH, Incheon Sejong Hospital; CUIMC, Columbia University Irving Medical Center.

Composite AI-ECG–guided selection reduced the number of echocardiograms needed to identify one SHD case from approximately 7.9 to 2.1 in ISH, with directionally consistent gains in CUIMC; the magnitude depends on underlying prevalence and is not directly comparable across cohorts. Across subgroups, composite performance was generally consistent, with lower specificity in atrial fibrillation, prior HF, chronic kidney disease, LBBB, and pacemaker rhythm (Supplementary Table 6).

### Prediction of new-onset SHD

Among individuals without baseline SHD or HF, 4,110 were included in ISH (median follow-up, 2.9 years; IQR, 1.7–4.5), of whom 390 (9.5%) developed new-onset SHD—incident reduced LVEF in 202 (4.9%), VHD in 103 (2.5%), LVH in 35 (0.9%), and PH in 123 (3.0%).

In UKB, 41,226 individuals were at risk (median follow-up, 3.3 years; IQR, 2.4–4.8), of whom 783 (1.9%) developed new-onset SHD—incident reduced LVEF in 234 (0.6%), VHD in 326 (0.8%), LVH in 325 (0.8%), and PH in 32 (0.1%).

Baseline composite AI-ECG positivity was significantly associated with new-onset SHD in both cohorts (Table 2). In ISH, composite-positive individuals had an approximately 3.8-fold higher risk than composite-negative individuals (HR, 3.75; 95% CI, 2.99–4.71), with consistent risk elevation across individual phenotypes and discrimination (C statistic) ranging from 0.69 to 0.78. In UKB, composite positivity was similarly associated with new-onset SHD (HR, 2.75; 95% CI, 2.37–3.19), with directionally consistent associations across phenotypes and comparable discrimination (C statistic, 0.72–0.78).

**Table 2.** New-onset SHD Outcomes According to Baseline AI-ECG composite.

| Outcome (new-onset) | HR (95% CI) | C-statistic |
| --- | --- | --- |
| **ISH** |  |  |
| New-onset SHD (Composite) | 3.75 (2.99–4.71) | 0.72 (0.69–0.75) |
| New-onset Reduced LVEF | 4.06 (2.96–5.56) | 0.70 (0.65–0.74) |
| New-onset VHD | 4.08 (2.63–6.34) | 0.76 (0.70–0.82) |
| New-onset LVH | 8.67 (4.25–17.68) | 0.75 (0.64–0.85) |
| New-onset PH | 3.14 (2.10–4.69) | 0.78 (0.73–0.83) |
| **UKB** |  |  |
| New-onset SHD (Composite) | 2.75 (2.37–3.19) | 0.73 (0.71–0.75) |
| New-onset HF | 3.75 (2.87–4.88) | 0.78 (0.75–0.81) |
| New-onset VHD | 2.25 (1.78–2.83) | 0.72 (0.69–0.75) |
| New-onset LVH | 2.74 (2.17–3.46) | 0.72 (0.69–0.75) |
| New-onset PH | 2.71 (1.32– 5.57) | 0.78 (0.69–0.86) |

Kaplan–Meier analyses showed clear and consistent separation in the cumulative incidence of new-onset SHD by baseline composite status in both cohorts (Figure 3). Composite-positive individuals had a significantly higher cumulative incidence than composite-negative individuals (log-rank P < .001), with similar separation for individual components, including reduced LVEF, VHD, LVH, and PH (Supplementary Figure 8).

**Figure 3.**
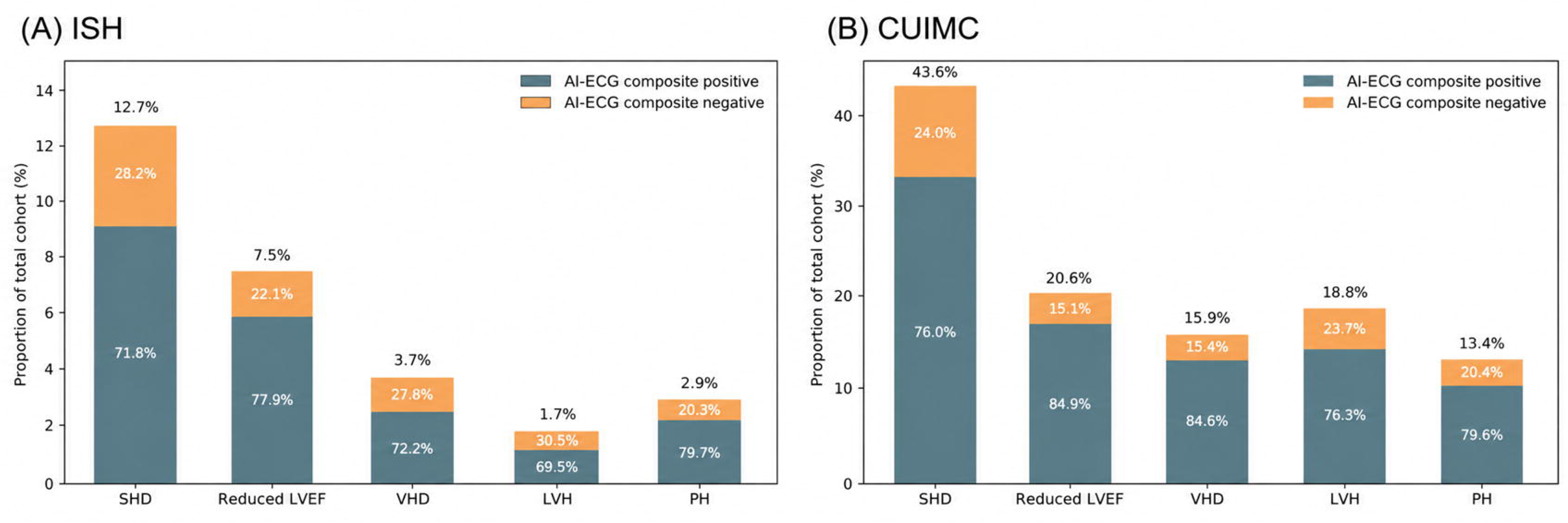
Prediction of new-onset SHD by baseline AI-ECG composite status Kaplan–Meier curves illustrate the cumulative incidence of new-onset composite SHD among individuals without baseline SHD or heart failure, stratified by baseline AI-ECG composite status. In both the ISH and UK Biobank cohorts, individuals classified as AI-ECG composite positive at baseline exhibited a significantly higher cumulative incidence of new-onset SHD over the follow-up period compared with those classified as composite negative (log-rank p < 0.0001). Shaded areas represent 95% confidence intervals, and the numbers at-risk at each time point are displayed below the plots. AI-ECG composite positivity was defined as a positive result on either the AI-ECG model for LVSD or the AI-ECG model for LVDD. AI-ECG, artificial intelligence–enabled electrocardiogram; SHD, structural heart disease; LVSD, left ventricular systolic dysfunction; LVDD, left ventricular diastolic dysfunction; ISH, Incheon Sejong Hospital; UKB, UK Biobank.

## Discussion

In this study, a composite AI-ECG combining LVSD and LVDD models identified prevalent SHD and predicted incident SHD across three multinational cohorts. Detection was consistent across diverse phenotypes—reduced LVEF, moderate or severe VHD, LVH, and PH—in both ISH and CUIMC, and in an at-risk population, baseline composite positivity was associated with substantially increased risk of incident SHD in both ISH and the UK Biobank. These findings extend evidence that AI-ECG captures cardiovascular signals broader than their nominal training targets to the SHD spectrum, using an independent, clinically deployed model family across multinational populations.(11,12) (**Central Illustration**)

### Composite AI-ECG and the SHD spectrum

SHD has traditionally been categorized into distinct diagnostic entities—reduced LVEF, VHD, LVH, and PH.(1,2) Recent frameworks, however, increasingly conceptualize SHD and HF as a continuum extending from subclinical alterations to overt disease, within which AI-ECG signals function as digital biomarkers reflecting a broader cardiovascular state rather than markers of single diagnostic entities.(6,24) The consistent detection of multiple SHD phenotypes by the composite in this study is concordant with this framing and with evidence that AI-ECG models trained for specific conditions capture signals extending beyond their nominal training targets.(11)

Several patterns support interpreting the composite as a graded screening signal rather than a phenotype-specific classifier. The incomplete overlap between LVSD and LVDD positivity across phenotypes indicates that these signals carry both shared and phenotype-specific information, with LVDD-predominant patterns in phenotypes other than reduced LVEF. Composite positivity increased stepwise with the number of co-occurring SHD components, and individuals negative for both models exhibited a milder disease profile despite the presence of SHD—suggesting that AI-ECG negativity corresponds to a lower-burden or earlier-stage state rather than a failure of detection. Consistent with this, regions of simultaneously elevated LVSD and LVDD scores showed the highest prevalence of composite SHD.

These findings extend recent composite AI-ECG approaches. Whereas models such as rECHOmmend, EchoNext, and PRESENT-SHD aggregate disease-specific labels into a single algorithm to broaden detection yield,LL¹L we instead combined two clinically deployed LVSD and LVDD models at predefined thresholds—an approach that parallels work showing the same architecture independently predicts incident HF in US population-based cohorts.(8–10,12)

### AI-ECG and the SHD progression continuum

A key strength of this study is the evaluation of the composite’s prognostic role in an at-risk population without baseline SHD or prior HF. In this group, composite positivity was significantly associated with a higher risk of developing diverse forms of SHD—not a single subtype—suggesting that the signal identifies a preclinical state with greater risk of progression to overt SHD, rather than merely reflecting disease missed by baseline imaging.(9,12) This indicates that the composite captures individuals on a trajectory of SHD progression, sensitive to the cumulative burden of disease over time.

Importantly, AI-ECG composite positivity was consistently associated with new SHD across multiple phenotypes, not just a single subtype. This indicates that the AI-ECG composite identifies individuals on a trajectory of SHD progression, reflecting sensitivity to the cumulative burden of SHD over time.

This interpretation aligns with HF frameworks that view heart failure as a continuous process encompassing preclinical stages.(1,2,24) Diastolic dysfunction and myocardial remodeling are recognized components of the preclinical SHD spectrum, and LVDD-based AI-ECG signals may precede overt structural change and clinically relevant elevations in filling pressure.(25) Together, these findings support viewing SHD and HF as a spectrum and position the composite as an upstream risk marker.

### Clinical implications and deployment positioning

These findings suggest that the composite AI-ECG can identify both current SHD and individuals at increased risk of progression, and may therefore complement existing screening by enabling more efficient identification of high-risk groups and targeted referral for further evaluation or preventive management.(12,26,27) A key practical advantage is its direct deployability: both the LVSD and LVDD models have undergone rigorous clinical validation and are actively deployed in practice, providing binary outputs at predefined thresholds, with the systolic model showing robust performance across multinational external validation cohorts, including resource-limited settings.(19,20,22,28,29) By leveraging these outputs without retraining or recalibration, the composite can be implemented within existing workflows without access to raw probabilities or institution-specific threshold adjustment—a deliberate design reflecting how AI-ECG tools are actually used at the point of care.

This binary composite framework is reinforced by convergent external evidence. Desai et al. recently showed that an identical strategy—combining systolic and diastolic dysfunction models by an OR rule at predefined thresholds—robustly predicted incident HF across three US population-based cohorts, with composite-positive individuals facing more than 20-fold higher near-term HF risk.(12) The convergence of these independent findings, across model families and clinical outcomes, supports the binary composite as both a principled and immediately actionable instantiation of AI-ECG as a broad cardiovascular biomarker.

The composite is also architecturally distinct from approaches that train a single end-to-end model on aggregated SHD labels.(8–10) When compared with a separately developed end-to-end SHD model trained on structural labels from the same development cohort, the composite achieved comparable performance without clinically meaningful loss of detection (Supplementary Figure 2 and Supplementary Table 4), supporting it as a viable architectural alternative. Calibration of a continuous composite score, derived from the average of the LVSD and LVDD outputs, showed reasonable agreement between predicted and observed SHD prevalence in both cohorts (Supplementary Figure 9). The composite is particularly relevant where access to cardiac imaging and specialist care is limited, positioning it not as a diagnostic tool but as a deployable, scalable cardiovascular screening strategy.(30)

### Limitations

This study has several limitations. First, the observational design precludes causal inference, and whether AI-ECG–based risk stratification improves outcomes through targeted intervention requires prospective validation. Second, incident SHD was not ascertained on a protocol-driven schedule: echocardiography was performed on clinical indication in ISH, and incident events were identified from ICD diagnosis codes in the UK Biobank. These approaches may introduce surveillance bias and outcome misclassification, particularly for milder or subclinical phenotypes, and prospective studies with standardized imaging follow-up are needed. Third, although the composite consistently detected phenotypes other than reduced LVEF, we did not formally test whether this detection is independent of coexisting reduced LVEF, which merits dedicated investigation. Finally, SHD was defined using dichotomized echocardiographic criteria and the models were applied at predefined thresholds; the composite should therefore be interpreted as a screening signal rather than a quantitative measure of disease severity or stage.

## Conclusions

A composite AI-ECG combining independent, clinically deployed LVSD and LVDD models identified prevalent and predicted incident SHD across three multinational cohorts, with consistent associations across diverse phenotypes and populations. Applied without retraining, it offers an immediately actionable, scalable approach to structural heart disease screening beyond its individual training targets.

## PERSPECTIVES

### COMPETENCY IN PATIENT CARE AND PROCEDURAL SKILLS

A composite AI-ECG that combines independently developed, clinically deployed models for left ventricular systolic and diastolic dysfunction identifies individuals with prevalent structural heart disease across multiple phenotypes—and those at increased risk of incident disease—using only the standard 12-lead ECG. Because it operates on binary model outputs without retraining or recalibration, it can be embedded within existing workflows as a scalable screen to prioritize echocardiographic referral, particularly where access to cardiac imaging and specialist evaluation is limited.

## TRANSLATIONAL OUTLOOK

Whether AI-ECG–guided screening improves diagnostic yield and downstream clinical outcomes remains unproven and requires prospective, ideally randomized, evaluation. Further work should test the composite against protocol-driven imaging follow-up to reduce surveillance bias, clarify its detection of phenotypes beyond reduced ejection fraction independent of coexisting systolic dysfunction, and establish whether serial composite results can track progression along the structural heart disease continuum to guide the timing of preventive intervention.

## Abbreviations

AI-ECG: Artificial Intelligence–Enabled Electrocardiogram
ISH: Incheon Sejong Hospital
LVDD: Left Ventricular Diastolic
Dysfunction LVEF: Left Ventricular Ejection Fraction
LVH: Left Ventricular Hypertrophy
LVSD: Left Ventricular Systolic Dysfunction
PH: Pulmonary Hypertension
SHD: Structural Heart Disease
UKB: UK Biobank
VHD: Valvular Heart Disease

## Data Availability

The de-identified electrocardiogram dataset from Columbia University Irving Medical Center analyzed in this study was publicly released and is available as described in its original publication. UK Biobank data are available to approved researchers through the UK Biobank application process (https://www.ukbiobank.ac.uk); the present analyses were conducted under application 89372. The Incheon Sejong Hospital clinical data contain potentially identifying patient information and are not publicly available owing to institutional and patient-privacy restrictions; de-identified data may be made available from the corresponding author upon reasonable request, subject to approval by the Institutional Review Board of Incheon Sejong Hospital. The AI-ECG models evaluated in this study are proprietary and are not publicly available.

## Acknowledgments

None

---

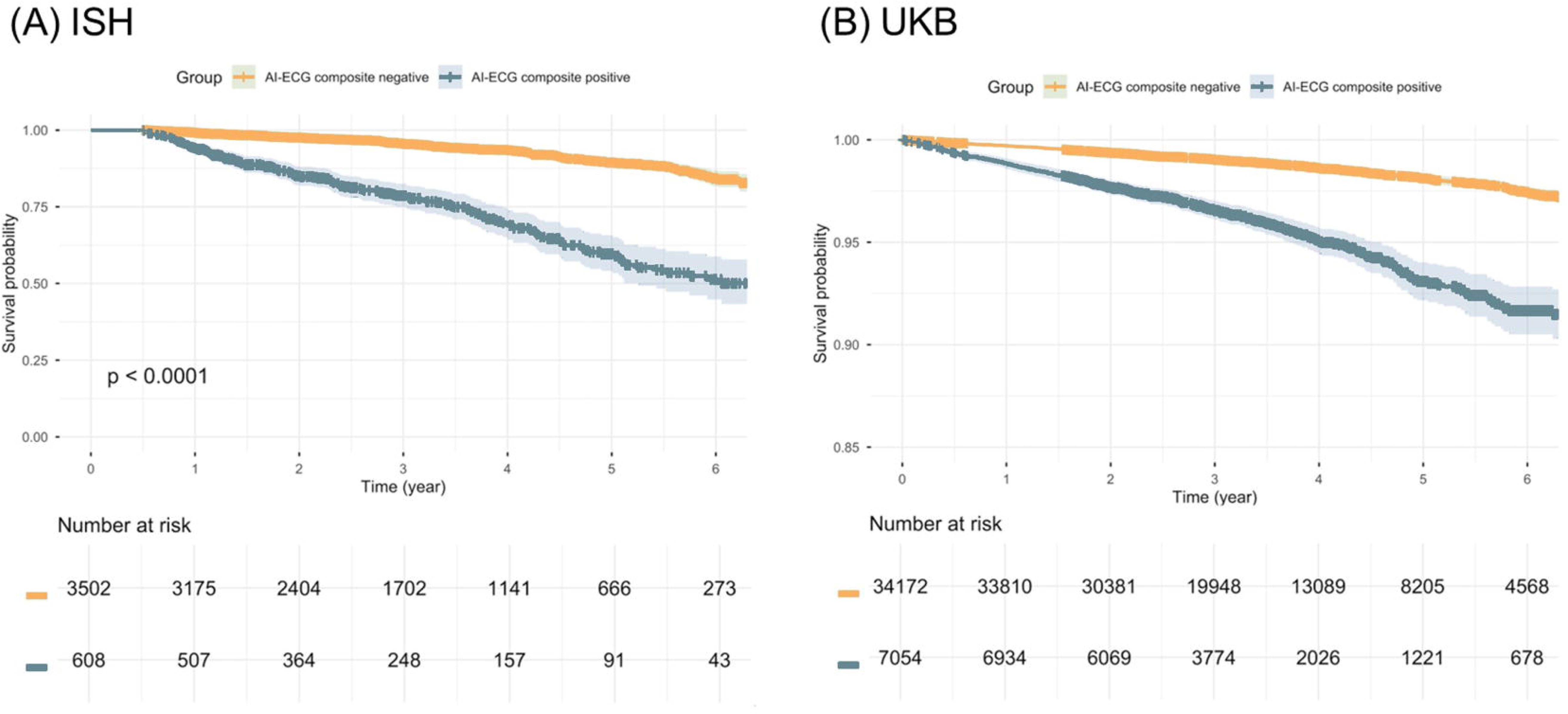
Central Illustration. Composite AI-ECG for Detection and Prediction of Structural Heart Disease Across Multinational Cohorts A composite artificial intelligence–enabled electrocardiogram (AI-ECG)—defined as a positive result on either an independently developed left ventricular systolic dysfunction (LVSD) model or a left ventricular diastolic dysfunction (LVDD) model—was evaluated across three multinational cohorts: Incheon Sejong Hospital (ISH; Republic of Korea), Columbia University Irving Medical Center (CUIMC; United States), and the UK Biobank (UKB; United Kingdom). In cross-sectional analyses (ISH and CUIMC), the composite detected prevalent structural heart disease (SHD)—reduced left ventricular ejection fraction, moderate or severe valvular heart disease, left ventricular hypertrophy, or pulmonary hypertension—with sensitivity of 71.8% and 76.1% and specificity of 88.3% and 70.1%, respectively. In longitudinal analyses of at-risk individuals without baseline SHD or heart failure (ISH and UKB), baseline composite positivity was associated with incident SHD (hazard ratios, 3.75 and 2.75), with consistent separation of cumulative-incidence curves by composite status. Applied without retraining or recalibration, the composite identified both prevalent and emerging SHD beyond its individual training targets, supporting its potential as a scalable cardiovascular screening tool. AI-ECG, artificial intelligence–enabled electrocardiogram; SHD, structural heart disease; LVSD, left ventricular systolic dysfunction; LVDD, left ventricular diastolic dysfunction; ISH, Incheon Sejong Hospital; CUIMC, Columbia University Irving Medical Center; UKB, UK Biobank.

## References

1. Heidenreich PA, Bozkurt B, Aguilar D et al. 2022 AHA/ACC/HFSA Guideline for the Management of Heart Failure: Executive Summary: A Report of the American College of Cardiology/American Heart Association Joint Committee on Clinical Practice Guidelines. J Am Coll Cardiol 2022;79:1757–1780.

2. McDonagh TA, Metra M, Adamo M et al. 2023 focused update of the 2021 ESC guidelines for the diagnosis and treatment of acute and chronic heart failure: developed by the task force for the diagnosis and treatment of acute and chronic heart failure of the European Society of Cardiology (ESC) with the special contribution of the Heart Failure Association (HFA) of the ESC. European heart journal 2023;44:3627–3639.

3. Alkhouli M, Alqahtani F, Holmes DR, Berzingi C. Racial disparities in the utilization and outcomes of structural heart disease interventions in the United States. Journal of the American Heart Association 2019;8:e012125.

4. Joynt Maddox KE, Elkind MS, Aparicio HJ et al. Forecasting the burden of cardiovascular disease and stroke in the United States through 2050—prevalence of risk factors and disease: a presidential advisory from the American Heart Association. Circulation 2024;150:e65–e88.

5. Tsao CW, Aday AW, Almarzooq ZI, et al. Heart disease and stroke statistics—2023 update: a report from the American Heart Association. Circulation 2023;147:e93–e621.

6. Lala A, Beavers C, Blumer V et al. The continuum of prevention and heart failure in cardiovascular medicine: a joint scientific statement from the Heart Failure Society of America and the American Society for Preventive Cardiology. Journal of cardiac failure 2025.

7. Elias P, Jain SS, Poterucha T et al. Artificial Intelligence for Cardiovascular Care-Part 1: Advances: JACC Review Topic of the Week. J Am Coll Cardiol 2024;83:2472–2486.

8. Poterucha TJ, Jing L, Ricart RP et al. Detecting structural heart disease from electrocardiograms using AI. Nature 2025;644:221–230.

9. Dhingra LS, Aminorroaya A, Sangha V et al. Ensemble deep learning algorithm for structural heart disease screening using electrocardiographic images: PRESENT SHD. Journal of the American College of Cardiology 2025;85:1302–1313.

10. Ulloa-Cerna AE, Jing L, Pfeifer JM et al. rECHOmmend: An ECG-Based Machine Learning Approach for Identifying Patients at Increased Risk of Undiagnosed Structural Heart Disease Detectable by Echocardiography. Circulation 2022;146:36–47.

11. Croon PM, Dhingra LS, Biswas D, Oikonomou EK, Khera R. Phenotypic Selectivity of Artificial Intelligence–Enhanced Electrocardiography in Cardiovascular Diagnosis and Risk Prediction. Circulation 2025;152:1282–1294.

12. Desai AS, Pandey A, Suratekar R et al. Predicting Heart Failure From 12-Lead ECGs Using AI: A HeartShare/AMP-HF Pooled Cohort Analysis. J Am Coll Cardiol 2025.

13. Palmer LJ. UK Biobank: bank on it. The Lancet 2007;369:1980–1982.

14. Raisi-Estabragh Z, Petersen SE. Cardiovascular research highlights from the UK Biobank: opportunities and challenges. Cardiovascular Research 2020;116:e12–e15.

15. Nagueh SF, Sanborn DY, Oh JK et al. Recommendations for the Evaluation of Left Ventricular Diastolic Function by Echocardiography and for Heart Failure With Preserved Ejection Fraction Diagnosis: An Update From the American Society of Echocardiography. J Am Soc Echocardiogr 2025;38:537–569.

16. Otto CM, Nishimura RA, Bonow RO et al. 2020 ACC/AHA Guideline for the Management of Patients With Valvular Heart Disease: A Report of the American College of Cardiology/American Heart Association Joint Committee on Clinical Practice Guidelines. Circulation 2021;143:e72–e227.

17. Praz F, Borger MA, Lanz J et al. 2025 ESC/EACTS Guidelines for the management of valvular heart disease. Eur Heart J 2025;46:4635–4736.

18. Kwon JM, Kim KH, Jeon KH et al. Development and Validation of Deep-Learning Algorithm for Electrocardiography-Based Heart Failure Identification. Korean Circ J 2019;49:629–639.

19. Jeong JH, Kang S, Lee HS et al. Deep learning algorithm for predicting left ventricular systolic dysfunction in atrial fibrillation with rapid ventricular response. Eur Heart J Digit Health 2024;5:683–691.

20. Lee HS, Lee S, Kang S et al. Artificial Intelligence–Enabled ECG Screening for LVSD in LBBB: Evaluating Model Development and Transfer Learning Approaches. JACC: Advances 2025;4:102089.

21. Lee MS, Jang J-H, Kang S et al. Transparent and robust Artificial intelligence-driven Electrocardiogram model for Left Ventricular Systolic Dysfunction. Diagnostics 2025;15:1837.

22. Lim J, Lee MS, Suh JH et al. Artificial IntelligenceLEnabled ECG for Elevated E/e’on Echocardiography: Hemodynamic Relevance and Prognostic Value. Journal of the American Heart Association 2025:e046989.

23. Song J, Jang J-H, Hong D, Kwon J-m, Jo Y-Y. CREMA: A Contrastive Regularized Masked Autoencoder for Robust ECG Diagnostics across Clinical Domains. arXiv preprint arXiv:240707110 2024.

24. Bozkurt B, Coats AJ, Tsutsui H et al. Universal definition and classification of heart failure: a report of the heart failure society of America, heart failure association of the European society of cardiology, Japanese heart failure society and writing committee of the universal definition of heart failure: endorsed by the Canadian heart failure society, heart failure association of India, cardiac society of Australia and New Zealand, and Chinese heart failure association. European Journal of Heart Failure 2021;23:352–380.

25. Lee E, Ito S, Miranda WR et al. Artificial intelligence-enabled ECG for left ventricular diastolic function and filling pressure. npj Digital Medicine 2024;7:4.

26. Lin C-S, Liu W-T, Tsai D-J et al. AI-enabled electrocardiography alert intervention and all-cause mortality: a pragmatic randomized clinical trial. Nature Medicine 2024;30:1461–1470.

27. Yao X, Rushlow DR, Inselman JW et al. Artificial intelligence-enabled electrocardiograms for identification of patients with low ejection fraction: a pragmatic, randomized clinical trial. Nat Med 2021;27:815–819.

28. Croon PM, Boonstra MJ, Allaart CP et al. Artificial Intelligence–Enhanced Electrocardiogram Models for Detection of Left Ventricular Dysfunction: A Comparison Study. JACC: Advances 2026;5:102572.

29. Pandey A, Keshvani N, Segar MW et al. Artificial Intelligence Electrocardiogram and Left Ventricular Systolic Dysfunction in Kenya. JAMA cardiology 2026.

30. Adedinsewo DA, Morales-Lara AC, Afolabi BB et al. Artificial intelligence guided screening for cardiomyopathies in an obstetric population: a pragmatic randomized clinical trial. Nature Medicine 2024;30:2897–2906.

