## Supplementary Material for "Composite Artificial Intelligence–Enabled Electrocardiogram for Detection and Prediction of Structural Heart Disease"

**Supplementary Online Content**

**Supplementary Methods**

**Supplementary Figure 1.** Derivation, Composite Definition, and External Validation of AI-ECG Models for SHD

**Supplementary 2.** Study Flow and Cohort Construction for the End-to-End AI-ECG SHD Model
**Supplementary 3.** Distribution of AI-ECG LVSD and LVDD Classification Patterns Across SHD Phenotypes
**Supplementary 4.** AI-ECG Model Positivity by Number of SHD Components

**Supplementary 5.** Echocardiographic Severity of Structural Heart Disease Across AI-ECG Phenotypes

**Supplementary 6.** Distribution of SHD Phenotypes Across the Joint AI-ECG LVSD and LVDD Score Space

**Supplementary 7.** Sensitivity of the AI-ECG Composite Model for Detection of Individual SHD Phenotypes
**Supplementary 8.** Cumulative Incidence of Individual SHD Components by Baseline AI-ECG Composite Status

**Supplementary 9.** Calibration of the AI-ECG composite score for detection of structural heart disease

**Supplementary Table 1.** Definitions of Structural Heart Disease Components Across Cohorts

**Supplementary Table 2.** TRIPOD+AI Checklist

**Supplementary Table 3.** Baseline Characteristics of Study Cohorts

**Supplementary Table 4.** Comparison of SHD Detection Performance Between the AI-ECG Composite and an End-to-End AI-ECG SHD Model at Matched Sensitivity

**Supplementary Table 5.** Performance of AI-ECG LVSD and LVDD Models Across SHD Phenotypes
**Supplementary Table 6**. Subgroup Analysis of AI-ECG Composite Performance for SHD Detection

**Supplementary Methods**

**1. Cohort Curation and ECG Acquisition**

In the ISH cohort, the most recent eligible ECG–transthoracic echocardiography pair was selected to harmonize eligibility with the publicly released CUIMC dataset. ECGs were acquired as standard 10-second, 12-lead recordings at 500 Hz using PAGEWRITER TC30 and TC70 devices (Philips) and stored in the MUSE Cardiology Information System (GE Healthcare). Demographic and baseline clinical characteristics were extracted from the electronic medical record. The CUIMC dataset was made available in conjunction with a prior study on AI-based SHD detection, with echocardiography-derived labels curated using standardized clinical criteria; demographic and clinical variables were obtained using the dataset's standardized definitions.

**2. AI-ECG Model Development**

Both models were derived using a two-stage framework based on a previously established ECG foundation model. The foundation model was pretrained on large-scale, heterogeneous unlabeled ECG data using a hybrid contrastive–generative self-supervised objective to learn generalizable ECG representations, then fine-tuned on task-specific labeled ECG–echocardiography datasets by adding a classification head and optimizing a binary cross-entropy loss. The AI-ECG LVSD model was trained to identify an LVEF ≤40%, and the AI-ECG LVDD model using an echocardiographic septal E/e′ ratio >15 as a surrogate marker; for both, ECGs lacking echocardiography within 14 days, missing essential measurements, or of poor signal quality were excluded.²¹,²² The continuous scores and predefined thresholds were applied directly to all study cohorts (**eFigure 1**).

To benchmark the binary composite against an end-to-end approach, an AI-ECG SHD detection model was additionally developed using the same foundation architecture and fine-tuning framework, trained directly on composite SHD labels rather than individual functional targets. For comparability, the same underlying datasets were used where possible; after applying identical exclusion criteria, 145,870 ECG–TTE pairs from 93,527 patients were included (**eFigure 2**) and randomly partitioned at the patient level into training (80%), validation (10%), and internal test (10%) sets, with model selection based on validation AUROC.

**3. Statistical Analysis (Extended)**

Subgroup analyses were performed across strata defined by age, sex, past medical history, and ECG rhythm (sinus rhythm, atrial fibrillation, left bundle branch block, right bundle branch block, left ventricular hypertrophy, and paced rhythm). For the exploratory joint-score analysis, AI-ECG LVSD and LVDD scores were binned in 10-point intervals (0–100), and the prevalence of SHD phenotypes was calculated within each bin and visualized as two-dimensional heatmaps.

**Supplementary Figure 1.** Derivation, composite definition, and external validation of AI-ECG models for SHD

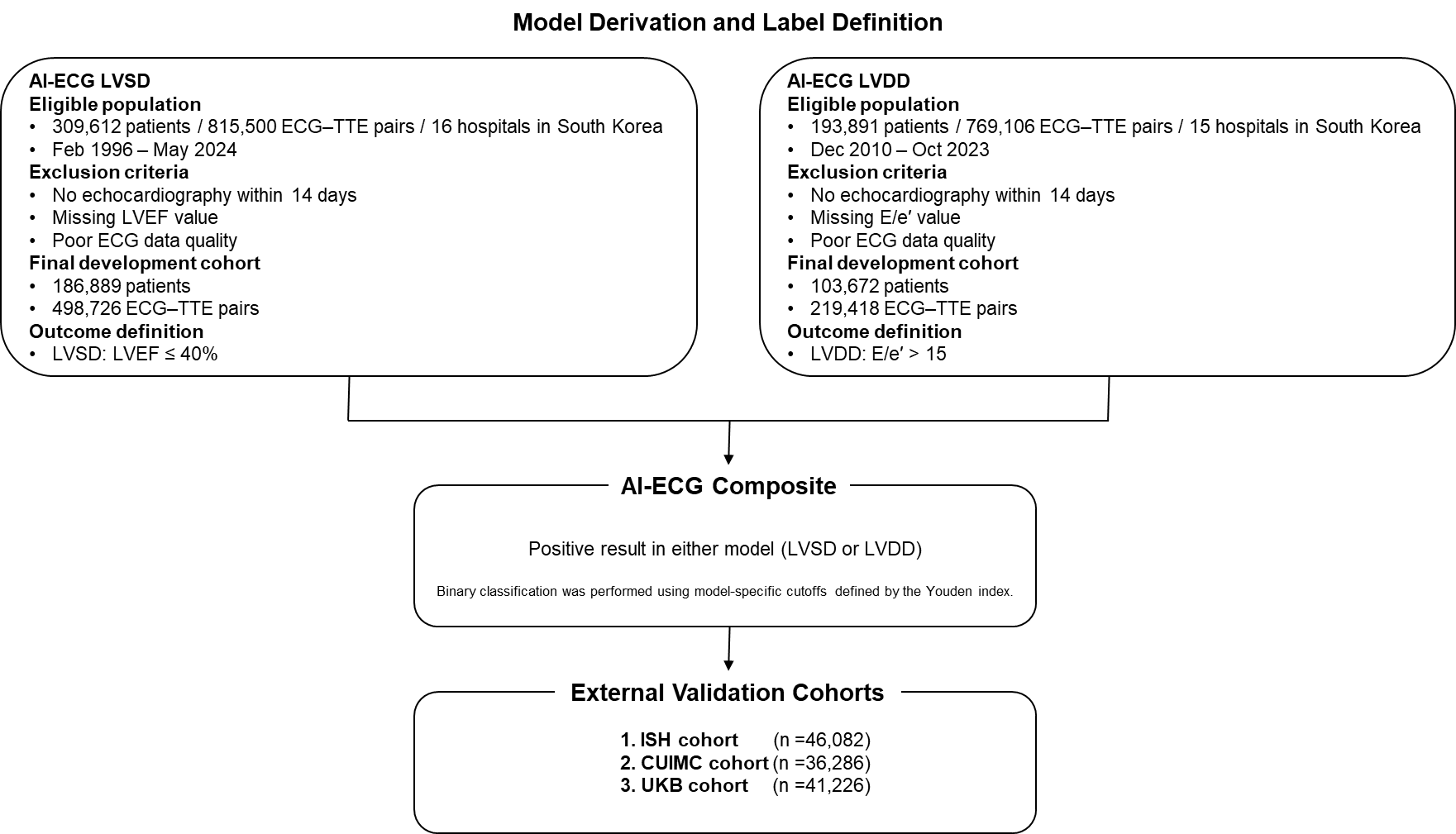

Overview of the derivation of AI-ECG models for LVSD and LVDD, the definition of the AI-ECG composite, and external validation cohorts. The AI-ECG LVSD model was derived using paired ECG–echocardiography data with LVSD defined as a LVEF ≤40%, and the AI-ECG LVDD model was derived using paired ECG–TTE data with LVDD defined as an E/e′ ratio >15. For both models, ECGs without echocardiography within 14 days, missing echocardiographic measurements, or poor ECG quality were excluded.
The AI-ECG composite was defined as a positive result on either the LVSD or LVDD model, with binary classification performed using model-specific cutoffs determined by the Youden index during model derivation. The composite was applied without retraining or recalibration and evaluated in three independent cohorts: ISH, CUIMC, and the UKB.

AI-ECG, artificial intelligence–enabled electrocardiography; LVSD, left ventricular systolic dysfunction; LVDD, left ventricular diastolic dysfunction; LVEF, left ventricular ejection fraction; TTE, transthoracic echocardiography; ECG, electrocardiogram; ISH, Incheon Sejong Hospital; CUIMC, Columbia University Irving Medical Center; UKB, UK Biobank.

**Supplementary Figure 2.** Study flow and cohort construction for AI-ECG SHD model development

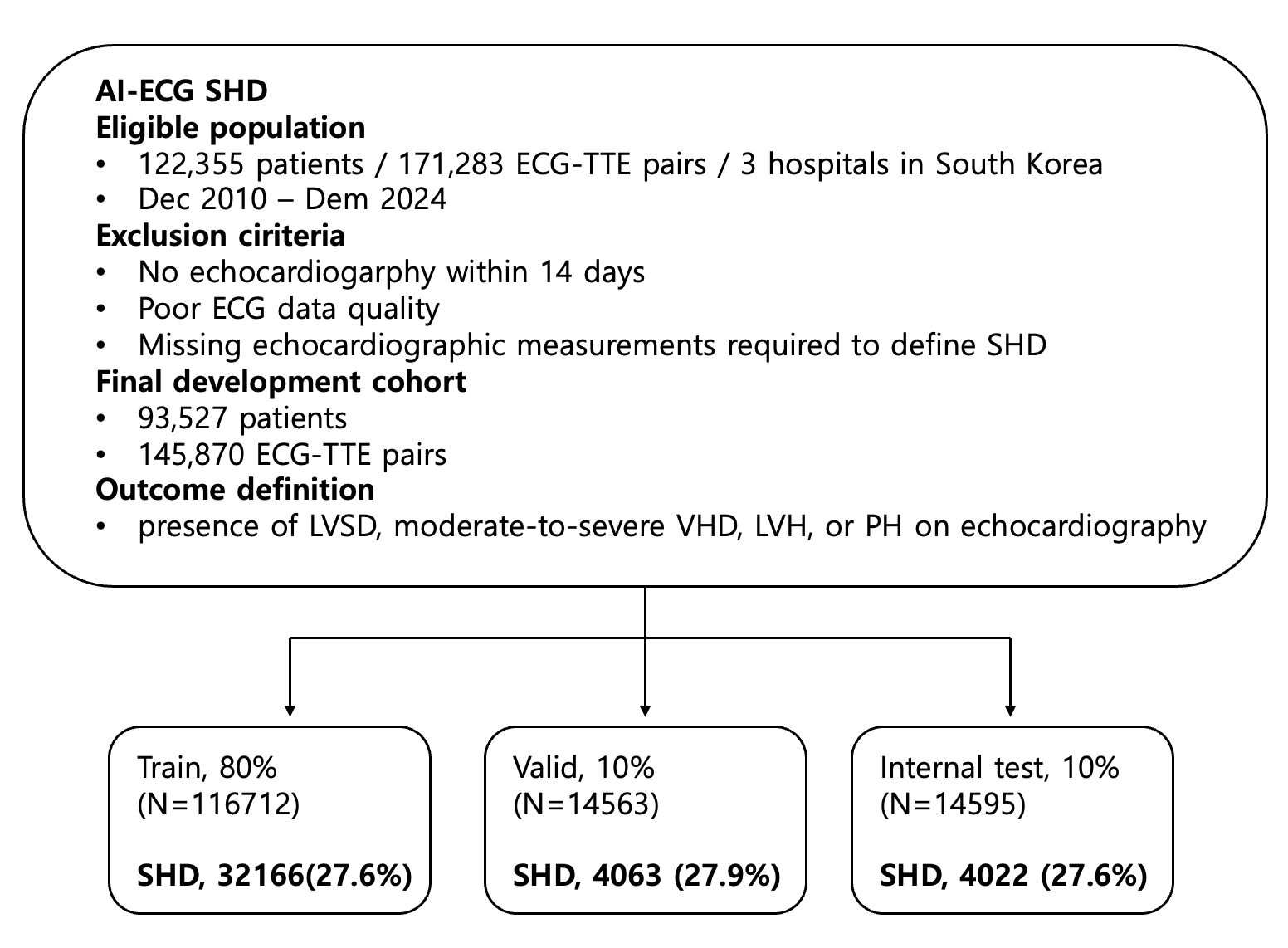

Flow diagram illustrating cohort construction and dataset partitioning for development of the AI-ECG SHD detection model. ECG–TTE pairs with sufficient echocardiographic information to ascertain SHD were included. SHD was defined according to predefined echocardiographic criteria, including reduced LVEF, VHD, LVH, or PH. ECGs without echocardiography within 14 days, missing required echocardiographic measurements, or poor ECG quality were excluded. The final dataset of 145,870 ECG–TTE pairs from 93,527 patients was randomly divided at the patient level into training (80%), validation (10%), and internal test (10%) sets.

AI-ECG, artificial intelligence–enabled electrocardiography; SHD, structural heart disease; ECG, electrocardiogram; TTE, transthoracic echocardiography; LVEF, left ventricular ejection fraction; VHD, valvular heart disease; LVH, left ventricular hypertrophy; PH, pulmonary hypertension.

**eFigure 3.** Distribution of AI-ECG LVSD and LVDD classification patterns across SHD phenotypes
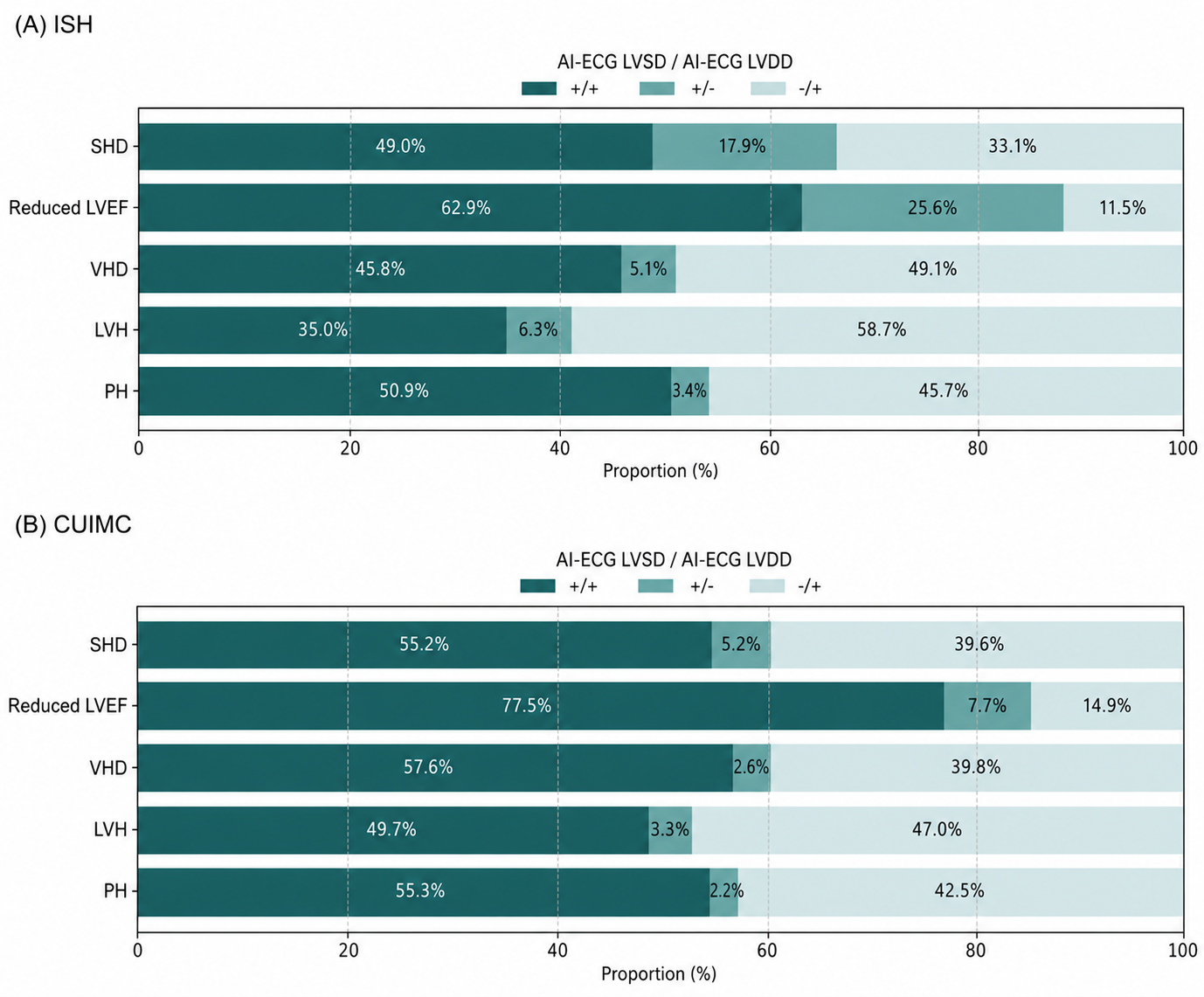

Horizontal stacked bar charts illustrate the distribution of joint AI-ECG LVSD and LVDD classification patterns across composite SHD and its individual components, including reduced LVEF, VHD, LVH, and PH, in the ISH and CUIMC cohorts. For each phenotype, bars are partitioned according to AI-ECG LVSD and LVDD results, categorized as positive for both models (+/+), positive for LVSD only (+/−), or positive for LVDD only (−/+). Percentages displayed within bars indicate the proportion of individuals within each phenotype classified into the corresponding AI-ECG LVSD/LVDD category.

AI-ECG, artificial intelligence–enabled electrocardiography; SHD, structural heart disease; LVSD, left ventricular systolic dysfunction; LVDD, left ventricular diastolic dysfunction; LVEF, left ventricular ejection fraction; VHD, valvular heart disease; LVH, left ventricular hypertrophy; PH, pulmonary hypertension; ISH, Incheon Sejong Hospital; CUIMC, Columbia University Irving Medical Center.

**Supplementary Figure 4.** AI-ECG model positivity by number of
SHD components in the ISH and CUIMC cohort

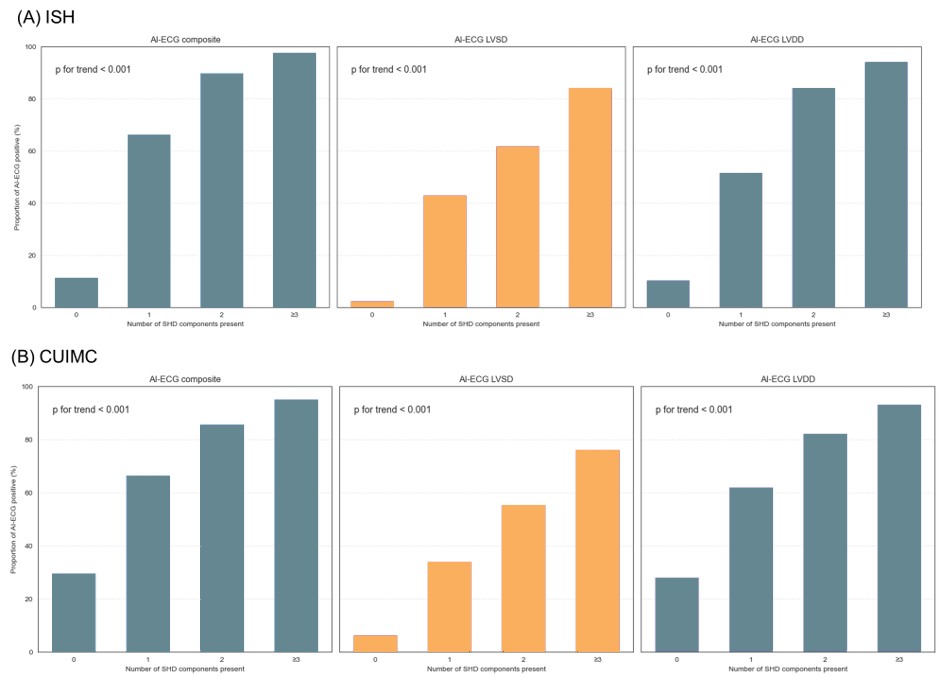

Bar plots show the proportion of participants with positive AI-ECG results across categories defined by the number of SHD components present (0, 1, 2, or ≥3). Panels display results for the AI-ECG composite, AI-ECG LVSD, and AI-ECG LVDD models. In each panel, percentages represent the proportion of participants within each SHD component category who were classified as positive by the corresponding AI-ECG model, demonstrating a graded increase in AI-ECG positivity with increasing SHD component burden. P values for trend were calculated using the Cochran–Armitage trend test.

AI-ECG, artificial intelligence–enabled electrocardiography; SHD, structural heart disease; ISH, Incheon Sejong Hospital; CUIMC, Columbia University Irving Medical Center.

**Supplementary Figure 5.** Echocardiographic severity of structural heart disease across AI-ECG phenotypes

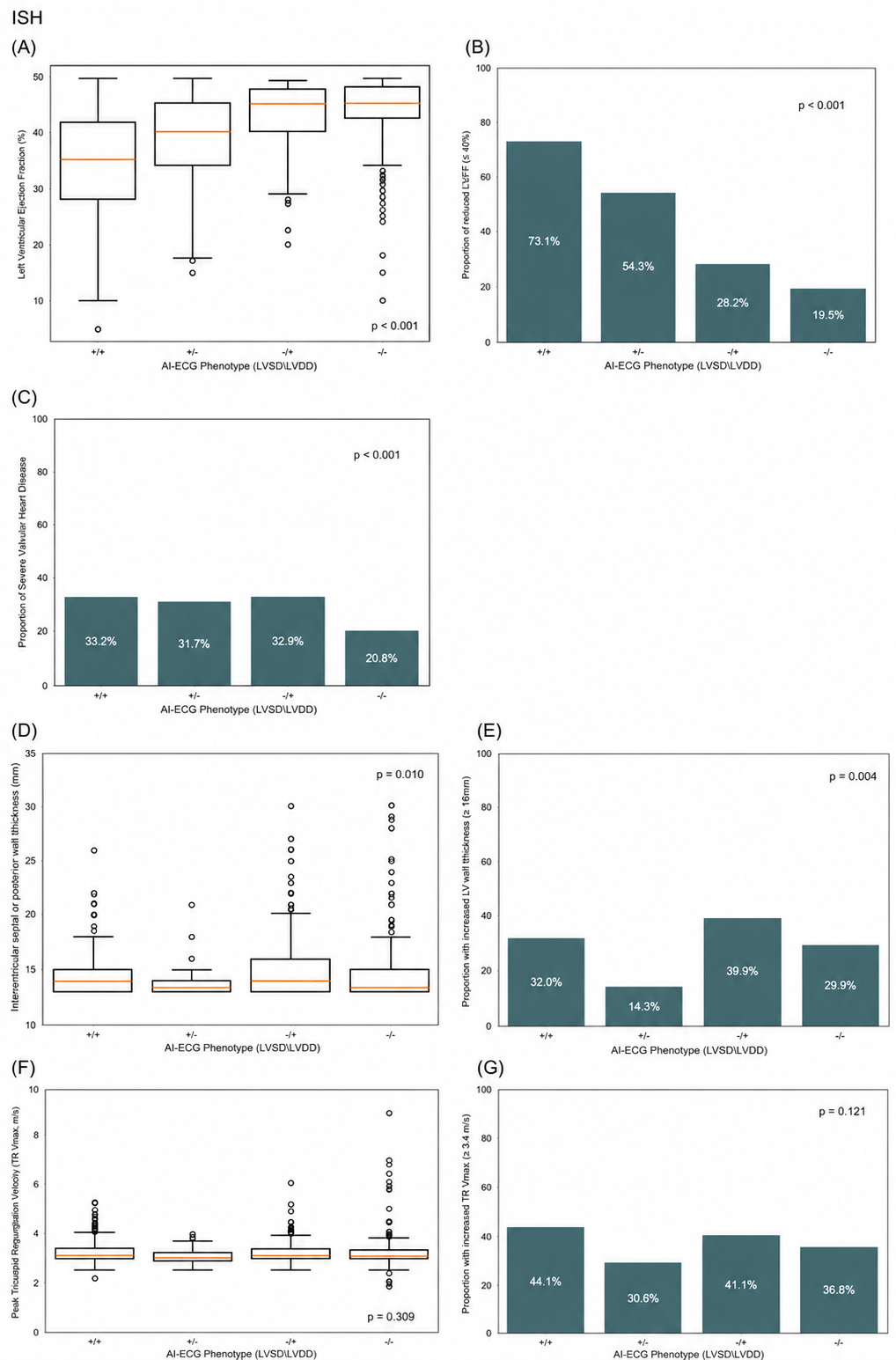

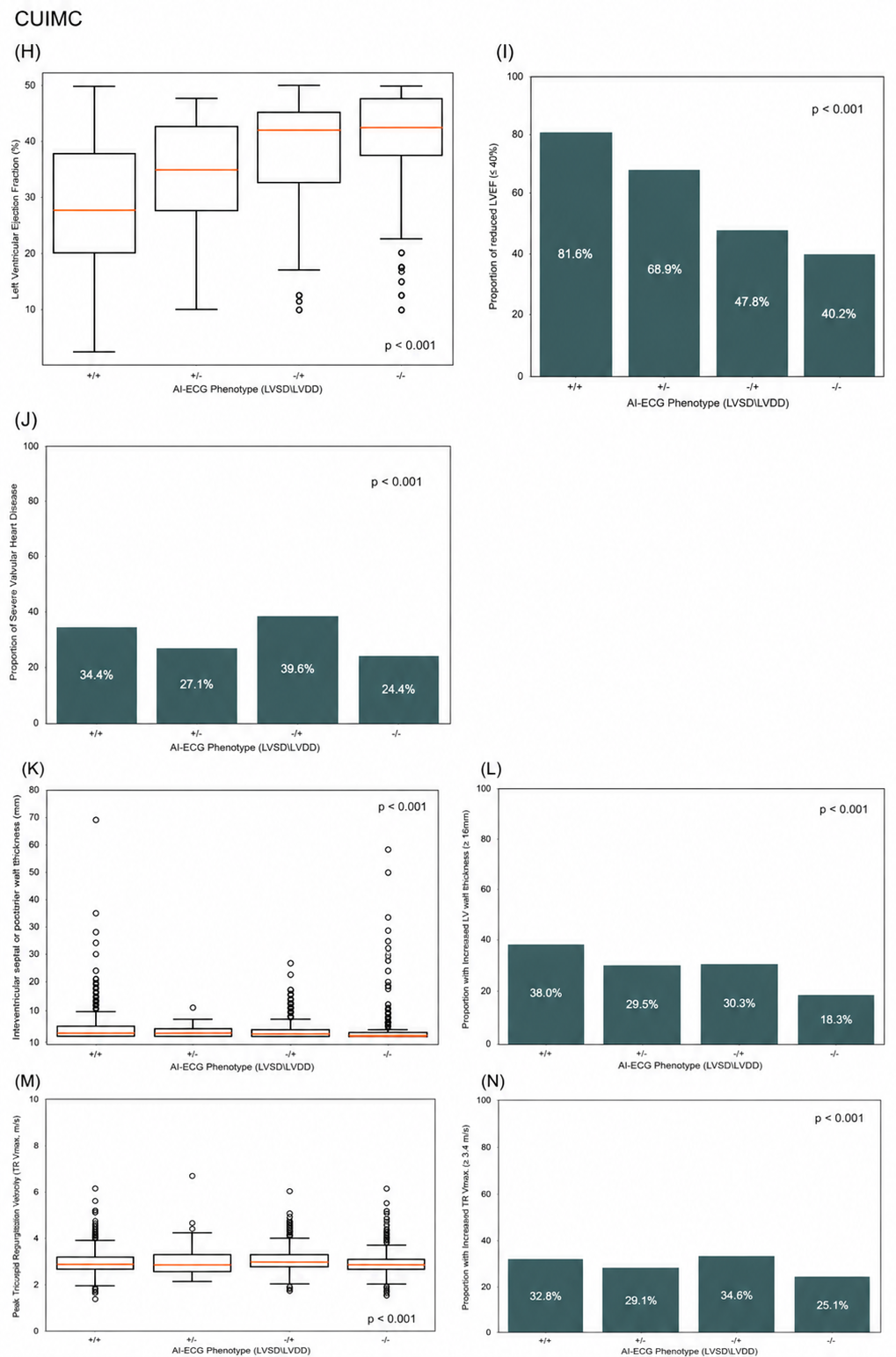

This figure illustrates differences in echocardiographic measures of SHD severity across AI-ECG phenotypes defined by the combined results of the LVSD and LVDD models (+/+, +/−, −/+, −/−). Panels display both continuous echocardiographic parameters (shown as box plots with median and interquartile range) and the corresponding proportions exceeding clinically relevant thresholds (shown as bar plots). Measures include LVEF and the proportion of LVEF ≤40% for reduced LVEF, the proportion of severe VHD, interventricular septal or posterior wall thickness and the proportion ≥15 mm for LVH, and peak TR Vmax and the proportion ≥3.4 m/s for PH. Continuous variables were compared using the Kruskal–Wallis test, and categorical variables were compared using the chi-square test. These findings demonstrate graded associations between AI-ECG phenotypes and echocardiographic markers of structural and hemodynamic disease severity.

AI-ECG, artificial intelligence–enabled electrocardiography; SHD, structural heart disease; LVSD, left ventricular systolic dysfunction; LVDD, left ventricular diastolic dysfunction; LVEF, left ventricular ejection fraction; VHD, valvular heart disease; LVH, left ventricular hypertrophy; PH, pulmonary hypertension; TR, tricuspid regurgitation; ISH, Incheon Sejong Hospital; CUIMC, Columbia University Irving Medical Center.

**Supplementary Figure 6.** Distribution of SHD phenotypes across the joint AI-ECG LVSD and LVDD score space

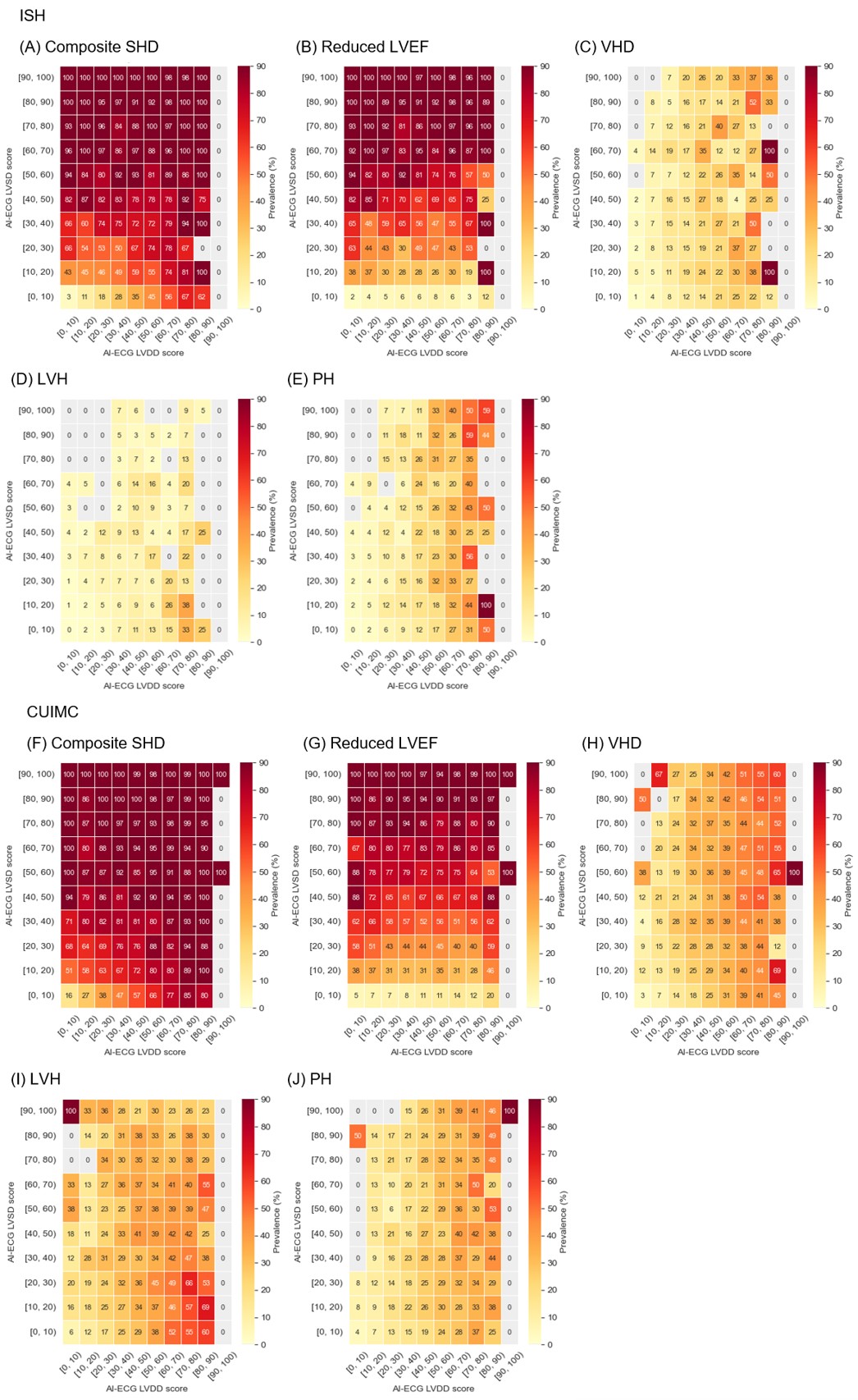

Heatmaps illustrating the prevalence of SHD phenotypes across the joint distribution of AI-ECG LVSD and LVDD scores in the ISH and CUIMC cohorts. AI-ECG LVDD scores are shown on the x-axis and AI-ECG LVSD scores on the y-axis. Scores were grouped in 10-point intervals (0–100) to generate a 10×10 grid. Within each bin, the prevalence of SHD phenotypes was calculated as the proportion of individuals with the corresponding condition among all individuals within that bin. Panels (A–E) present results from the ISH cohort and panels (F–J) present results from the CUIMC cohort. Heatmaps are shown for composite SHD, LVSD, VHD, LVH, and PH. Color intensity represents increasing prevalence, and bins without observations are shown in gray.

AI-ECG, artificial intelligence–enabled electrocardiography; SHD, structural heart disease; LVSD, left ventricular systolic dysfunction; LVDD, left ventricular diastolic dysfunction;
VHD, valvular heart disease; LVH, left ventricular hypertrophy; PH, pulmonary hypertension; ISH, Incheon Sejong Hospital; CUIMC, Columbia University Irving Medical Center.

**Supplementary Figure 7** Sensitivity of the AI-ECG Composite Model for Detection of Individual SHD Phenotypes

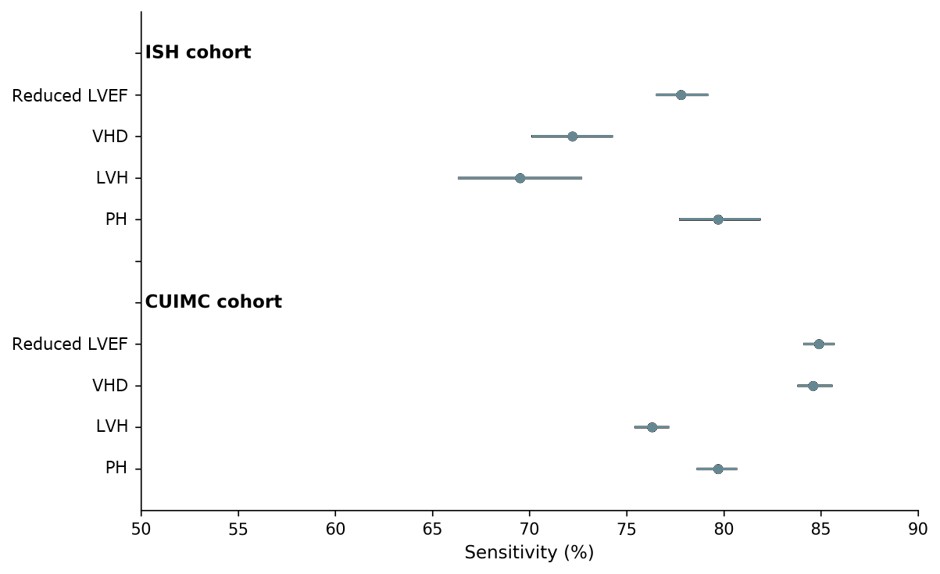

Forest plots showing the sensitivity (point estimates and 95% confidence intervals) of the AI-ECG composite model for detecting individual SHD phenotypes in the ISH and CUIMC cohorts. Sensitivity estimates are presented for reduced LVEF, VHD, LVH, and PH at the predefined thresholds.

AI-ECG, artificial intelligence–enabled electrocardiography; SHD, structural heart disease; LVSD, left ventricular systolic dysfunction; LVDD, left ventricular diastolic dysfunction; LVEF, left ventricular ejection fraction; VHD, valvular heart disease; LVH, left ventricular hypertrophy; PH, pulmonary hypertension; ISH, Incheon Sejong Hospital; CUIMC, Columbia University Irving Medical Center.

**Supplementary Figure 8.** Cumulative incidence of individual SHD components by baseline AI-ECG composite status

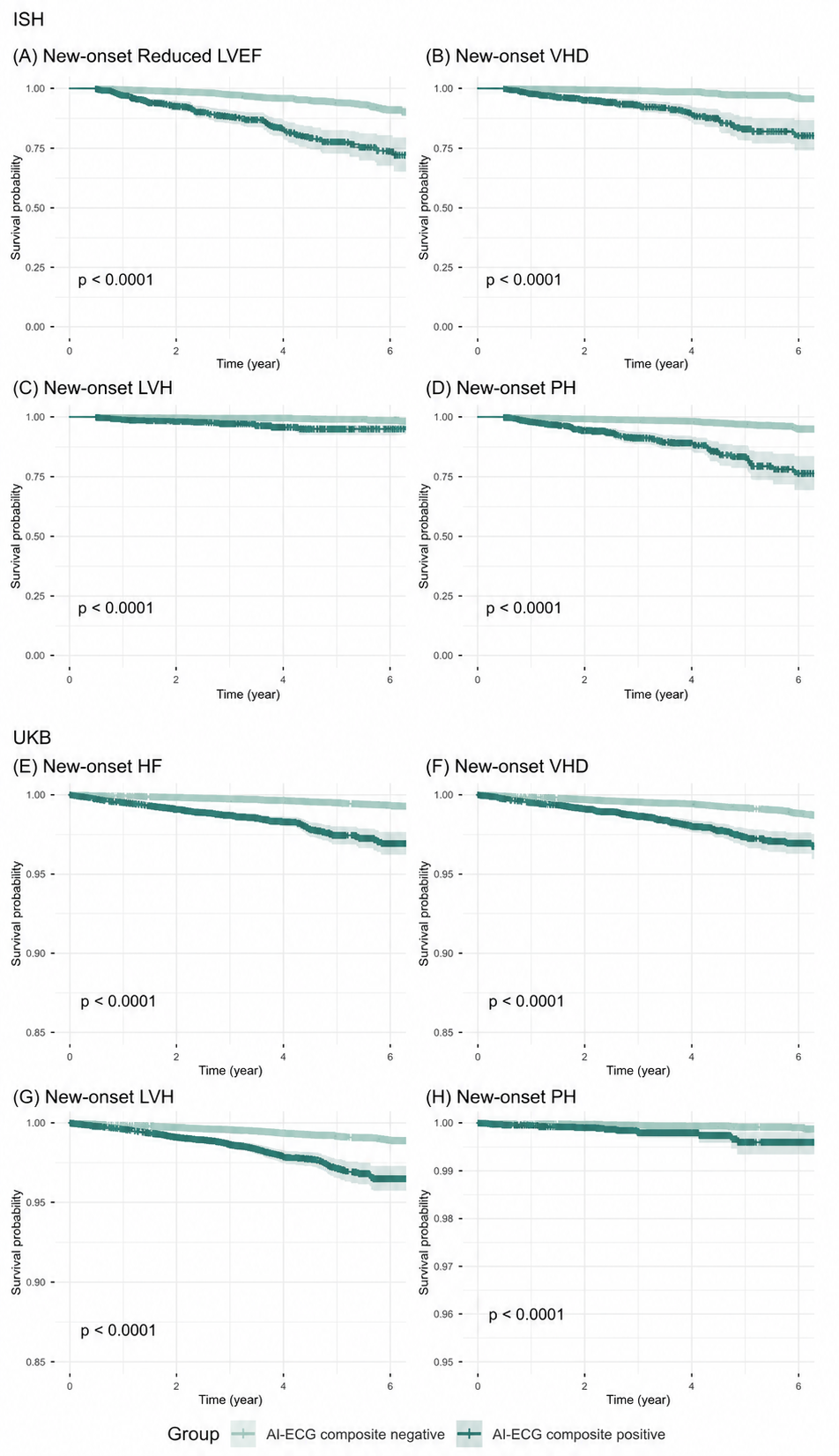

Kaplan–Meier curves showing the cumulative incidence of individual components of SHD among individuals without baseline SHD or heart failure, stratified by baseline AI-ECG composite status. Panels A–D correspond to the Incheon Sejong Hospital (ISH) cohort and depict new-onset (A) Reduced LVEF, (B) moderate or severe VHD, (C) LVH, and (D) PH. Panels E–H correspond to the UKB cohort and depict new-onset (E) Reduced LVEF, (F) moderate or severe VHD, (G) LVH, and (H) PH. Lighter teal lines indicate AI-ECG composite–negative individuals, and darker teal lines indicate AI-ECG composite–positive individuals.

In both the ISH and UK Biobank cohorts, individuals classified as AI-ECG composite positive at baseline demonstrated a consistently higher cumulative incidence of each individual SHD component compared with composite-negative individuals (P < .001). Shaded areas represent 95% confidence intervals, and numbers at risk are shown below each plot. AI-ECG composite positivity was defined as a positive result on either the AI-ECG model for left ventricular systolic dysfunction or the AI-ECG model for left ventricular diastolic dysfunction.

AI-ECG, artificial intelligence–enabled electrocardiography; SHD, structural heart disease; LVSD, left ventricular systolic dysfunction; LVEF, left ventricular ejection fraction; VHD, valvular heart disease; LVH, left ventricular hypertrophy; PH, pulmonary hypertension; ISH, Incheon Sejong Hospital; UKB, UK Biobank.

**Supplementary Figure 9.** Calibration of the AI-ECG composite score for detection of structural heart disease

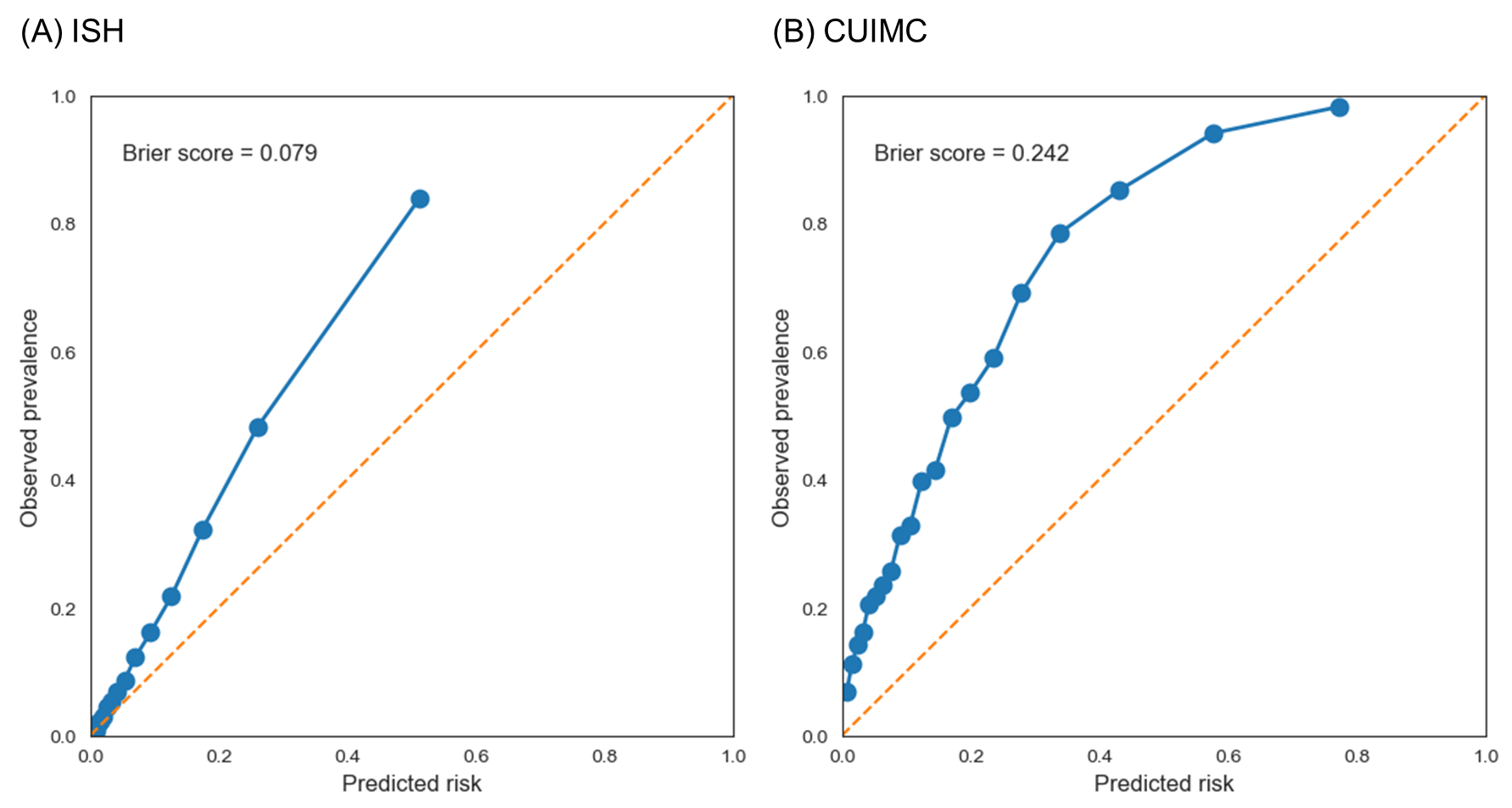

Calibration plots illustrating the relationship between predicted risk and observed prevalence of SHD in the ISH and CUIMC cohorts. The composite score was derived as the mean of the AI-ECG LVSD and LVDD probability outputs. Predicted risk was grouped using quantile-based binning, and the observed SHD prevalence was calculated within each bin. The dashed diagonal line represents perfect calibration. Brier scores are shown for each cohort.

AI-ECG, artificial intelligence–enabled electrocardiography; SHD, structural heart disease; LVSD, left ventricular systolic dysfunction; LVDD, left ventricular diastolic dysfunction; ISH, Incheon Sejong Hospital; CUIMC, Columbia University Irving Medical Center.

**Supplementary Table 1. Definitions of Structural Heart Disease Components Across Cohorts**

| **SHD component** | **Echocardiographic definition (ISH, CUIMC)** | **ICD-10 code (UKB)** |
| --- | --- | --- |
| Reduced LVEF | LVEF <50%ᵃ | I50ᵇ |
| Valvular heart disease | Moderate or severe aortic stenosis, aortic regurgitation, mitral stenosis, mitral regurgitation, or tricuspid regurgitation | I34, I35, I36, I05, I06, I07 |
| Left ventricular hypertrophy | Maximum LV wall thickness ≥13 mm^9^ | I51 |
| Pulmonary hypertension | TR jet maximum velocity ≥3.2 m/s^15^ | I27 |

CUIMC, Columbia University Irving Medical Center; ICD-10, International Classification of Diseases, Tenth Revision; ISH, Incheon Sejong Hospital; LV, left ventricular; LVEF, left ventricular ejection fraction; SHD, structural heart disease; TR, tricuspid regurgitation; UKB, UK Biobank.

ᵃ An LVEF of 41–49% corresponds to heart failure with mildly reduced ejection fraction.^¹,²^
ᵇ In UKB, echocardiography was unavailable; the systolic-dysfunction component was operationalized as incident heart failure (ICD-10 I50).

**Supplementary Table 2.** Transparent Reporting of a multivariable prediction model for Individual Prognosis Or Diagnosis + Artificial Intelligence (TRIPOD + AI) checklist.

Each item is addressed according to its location in the manuscript.

| **Section/Topic** | **Item** | **D/E** | **Checklist item (original wording)** | **Reported on page** |
| --- | --- | --- | --- | --- |
| TITLE | 1 | D;E | Identify the study as developing or evaluating the performance of a multivariable prediction model, the target population, and the outcome to be predicted | Title |
| ABSTRACT | 2 | D;E | See TRIPOD+AI for Abstracts checklist | Abstract |
| INTRODUCTION – Background | 3a | D;E | Explain the healthcare context (including whether diagnostic or prognostic) and rationale for developing or evaluating the prediction model, including references to existing models | Introduction |
|  | 3b | D;E | Describe the target population and the intended purpose of the prediction model in the context of the care pathway, including its intended users (e.g., healthcare professionals, patients, public) | Introduction |
|  | 3c | D;E | Describe any known health inequalities between sociodemographic groups | Introduction (multinational cohorts described) |
| INTRODUCTION – Objectives | 4 | D;E | Specify the study objectives, including whether the study describes the development or validation of a prediction model (or both) | Introduction (final paragraph) |
| METHODS – Data | 5a | D;E | Describe the sources of data separately for the development and evaluation datasets (e.g., randomised trial, cohort, routine care or registry data), the rationale for using these data, and representativeness of the data | Methods – Study Design and Data Sources |
|  | 5b | D;E | Specify the dates of the collected participant data, including start and end of participant accrual; and, if applicable, end of follow-up | Methods – Study Design |
| METHODS – Participants | 6a | D;E | Specify key elements of the study setting (e.g., primary care, secondary care, general population) including the number and location of centres | Methods – Study Design; Table 1 |
|  | 6b | D;E | Describe the eligibility criteria for study participants | Methods – Study Design |
|  | 6c | D;E | Give details of any treatments received, and how they were handled during model development or evaluation, if relevant | Not applicable |
| METHODS – Data preparation | 7 | D;E | Describe any data pre-processing and quality checking, including whether this was similar across relevant sociodemographic groups | Methods – AI-ECG Models and Composite |
| METHODS – Outcome | 8a | D;E | Clearly define the outcome that is being predicted and the time horizon, including how and when assessed, the rationale for choosing this outcome, and whether the method of outcome assessment is consistent across sociodemographic groups | Methods – Study Outcomes |
|  | 8b | D;E | If outcome assessment requires subjective interpretation, describe the qualifications and demographic characteristics of the outcome assessors | Methods – Study Outcomes |
|  | 8c | D;E | Report any actions to blind assessment of the outcome to be predicted | AI-ECG applied independently (implicit) |
| METHODS – Predictors | 9a | D | Describe the choice of initial predictors (e.g., literature, previous models, all available predictors) and any pre-selection of predictors before model building | Methods – AI-ECG models |
|  | 9b | D;E | Clearly define all predictors, including how and when they were measured (and any actions to blind assessment of predictors for the outcome and other predictors) | Methods – AI-ECG models |
|  | 9c | D;E | If predictor measurement requires subjective interpretation, describe the qualifications and demographic characteristics of the predictor assessors | Not applicable |
| METHODS – Sample size | 10 | D;E | Explain how the study size was arrived at (separately for development and evaluation), and justify that the study size was sufficient to answer the research question. Include details of any sample size calculation | Cohort sizes described; no formal calculation |
| METHODS – Missing data | 11 | D;E | Describe how missing data were handled. Provide reasons for omitting any data | Methods – Exclusion of missing ECG/TTE |
| METHODS – Analytical methods | 12a | D | Describe how the data were used (e.g., for development and evaluation of model performance) in the analysis, including whether the data were partitioned, considering any sample size requirements | Methods – AI-ECG Models and Composite |
|  | 12b | D | Depending on the type of model, describe how predictors were handled in the analyses (functional form, rescaling, transformation, or any standardisation) | Methods – AI-ECG Models and Composite |
|  | 12c | D | Specify the type of model, rationale, all model-building steps, including any hyperparameter tuning, and method for internal validation | Methods – AI-ECG Models and Composite |
|  | 12d | D;E | Describe if and how any heterogeneity in estimates of model parameter values and model performance was handled and quantified across clusters (e.g., hospitals, countries). See TRIPOD-Cluster for additional considerations | Subgroup analyses (Supplementary Table 2) |
|  | 12e | D;E | Specify all measures and plots used (and their rationale) to evaluate model performance (e.g., discrimination, calibration, clinical utility) and, if relevant, to compare multiple models | Statistical Analysis; Tables 2–3, Supplementary Tables 1, 3" |
|  | 12f | E | Describe any model updating (e.g., recalibration) arising from the model evaluation, either overall or for particular sociodemographic groups or settings | Not performed |
|  | 12g | E | For model evaluation, describe how the model predictions were calculated (e.g., formula, code, object, application programming interface) | Methods – AI-ECG models |
| METHODS – Class imbalance | 13 | D;E | If class imbalance methods were used, state why and how this was done, and any subsequent methods to recalibrate the model or the model predictions | Threshold determined by Youden index; no additional imbalance method |
| METHODS – Fairness | 14 | D;E | Describe any approaches that were used to address model fairness and their rationale | Subgroup analyses by age, sex, comorbidities, ECG rhythm |
| METHODS – Model output | 15 | D | Specify the output of the prediction model (e.g., probabilities, classification). Provide details and rationale for any classification and how the thresholds were identified | Methods – AI-ECG models (probability output; Youden cutoff) |

**Supplementary Table 3.** Baseline Characteristics of Study Cohorts

| **Characteristic** | **ISH cohort** | **CUIMC cohort** |
| --- | --- | --- |
| Number of participants | 46082 | 36286 |
| Age, years (mean ± SD / median [IQR]) | 59.9 ± 15.8 / 60.0 (50.0–71.0) | 62.5 ± 16.5 / 64.0 (52.0–75.0) |
| Female sex, n (%) | 22111 (48.0) | 18333 (50.5) |
| BMI, kg/m² (mean ± SD) | 24.7 ± 4.1 | - |
| Race, Ethnicity, n (%) |  |  |
| Asian | 46082 (100.0) | 1082 (3.0) |
| White | - | 10635 (29.3) |
| Black | - | 5424 (14.9) |
| Hispanic | - | 11006 (30.3) |
| Other | - | 2980 (8.2) |
| Unknown | - | 5159 (14.2) |
| **ECG feature (mean ± SD)** |  |  |
| Heart rate, bpm | 73.3 ± 16.0 | 80.4 ± 20.1 |
| PR interval | 167.5 ± 28.7 | 160.6 ± 32.3 |
| QRS duration | 97.1 ± 16.6 | 93.4 ± 20.9 |
| QT corrected | 436.8 ± 35.4 | 446.9 ± 37.3 |
| **ECG rhythm** |  |  |
| Normal sinus rhythm | 41931 (91.0) |  |
| Atrial fibrillation/atrial flutter | 3331 (7.2) | - |
| Left bundle branch block | 586 (1.3) | - |
| Right bundle branch block | 2122 (4.6) | - |
| Left ventricular hypertrophy | 10079 (21.9) | - |
| Pacemaker rhythm | 312 (0.7) | - |
| **Past medical history, n (%)** |  |  |
| Hypertension | 13466 (29.2) | - |
| Diabetes mellitus | 3919 (8.5) | - |
| Prior acute myocardial infarction | 1483 (3.2) | - |
| Prior heart failure | 8306 (18.0) | - |
| Atrial fibrillation | 4855 (10.5) | - |
| Chronic kidney disease | 1526 (3.3) | - |
| Stroke | 3801 (8.2) | - |
| **Echocardiographic parameters** |  |  |
| └ LVEF, % (mean ± SD) | 62.6 ± 9.1 | 53.4 ± 13.6 |
| **Structural heart disease categories, n (%)** |  |  |
| Composite of SHD | 5847 (12.7) | 15824 (43.6) |
| Reduced LVEF (<50%) | 3471 (7.5) | 7475 (20.6) |
| Moderate or severe VHD | 1696 (3.7) | 5755 (15.9) |
| Moderate or severe AS | 421 (0.9) | 1973 (5.4) |
| Moderate or severe AR | 275 (0.6) | 433 (1.2) |
| Moderate or severe MS | 144 (0.3) | - |
| Moderate or severe MR | 539 (1.2) | 2352 (6.5) |
| Moderate or severe TR | 626 (1.4) | 2534 (7.0) |
| LVH | 799 (1.7) | 6828 (18.8) |
| PH | 1341 (2.9) | 4850 (13.4) |

Values are presented as mean ± standard deviation, median (interquartile range), or number (percentage), as appropriate. Structural heart disease categories were defined based on echocardiographic assessments. Structural heart disease categories are not mutually exclusive. Missing data are indicated by dashes.

ISH, Incheon Sejong Hospital; CUIMC, Columbia University Irving Medical Center; BMI, body mass index; LVEF, left ventricular ejection fraction; SHD, structural heart disease; LVSD, left ventricular systolic dysfunction; AS, aortic stenosis; AR, aortic regurgitation; MS, mitral stenosis; MR, mitral regurgitation; TR, tricuspid regurgitation; LVH, left ventricular hypertrophy; PH, pulmonary hypertension.

**Supplementary Table 4**. Comparison of SHD detection performance between the AI-ECG composite and an AI-ECG SHD model at matched sensitivity

| **Cohort** | **Model** | **Sensitivity (%)** | **Specificity (%)** | **PPV (%)** | **NPV (%)** |
| --- | --- | --- | --- | --- | --- |
| ISH (n=46082) | AI-ECG composite | 71.8 (70.6–72.9) | 88.3 (88.0–88.6) | 47.2 (46.2–48.3) | 95.6 (95.4–95.8) |
|  | AI-ECG SHD | 71.8 (70.6–72.9) | 91.7 (91.5–92.0) | 55.8 (54.7–56.9) | 95.7 (95.5–95.9) |
| CUIMC (n=36286) | AI-ECG composite | 76.1 (75.4–76.7) | 70.1 (69.5–70.8) | 66.3 (65.6–67.0) | 79.1 (78.6–79.7) |
|  | AI-ECG SHD | 76.1 (75.5–76.8) | 73.8 (73.2–74.4) | 69.2 (68.6–69.9) | 80.0 (79.4–80.6) |

Performance comparison between the AI-ECG composite model and a separately developed AI-ECG model trained to detect SHD (AI-ECG SHD). To enable a fair comparison, the operating threshold of the AI-ECG SHD model was adjusted to match the sensitivity of the AI-ECG composite within each cohort. Specificity, PPV, and NPV were then compared at the matched sensitivity level. Performance metrics are shown with 95% confidence intervals.

ISH, Incheon Sejong Hospital; CUIMC, Columbia University Irving Medical Center; AI-ECG, artificial intelligence–enabled electrocardiography; SHD, structural heart disease; PPV, positive predictive value; NPV, negative predictive value.

**Supplementary Table 5.** Performance of AI-ECG LVSD and LVDD models across SHD phenotypes

| **SHD phenotype** | **Model** | **Sensitivity (%)** | **Specificity (%)** | **PPV (%)** | **NPV (%)** |
| --- | --- | --- | --- | --- | --- |
| ISH |  |  |  |  |  |
| Reduced LVEF | AI-ECG LVSD | 68.9 (67.3–70.4) | 96.5 (96.3–96.6) | 61.4 (59.8–62.9) | 97.4 (97.3–97.6) |
|  | AI-ECG LVDD | 57.9 (56.3–59.5) | 86.6 (86.3–87.0) | 26.1 (25.2–27.1) | 96.2 (96.0–96.4) |
|  | AI-ECG composite | 77.8 (76.5–79.2) | 85.5 (85.2–85.8) | 30.4 (29.5–31.4) | 97.9 (97.8–98.1) |
| VHD | AI-ECG LVSD | 36.8 (34.5–39.1) | 92.6 (92.4–92.9) | 16.0 (14.8–17.2) | 97.5 (97.3–97.6) |
|  | AI-ECG LVDD | 68.5 (66.4–70.7) | 85.3 (84.9–85.6) | 15.1 (14.3–15.9) | 98.6 (98.5–98.7) |
|  | AI-ECG composite | 72.2 (70.1–74.3) | 82.7 (82.4–83.1) | 13.8 (13.1–14.5) | 98.7 (98.6–98.8) |
| LVH | AI-ECG LVSD | 28.7 (25.4–31.7) | 91.9 (91.7–92.1) | 5.9 (5.1–6.6) | 98.6 (98.5–98.8) |
|  | AI-ECG LVDD | 65.2 (61.9–68.5) | 84.1 (83.8–84.5) | 6.8 (6.2–7.3) | 99.3 (99.2–99.4) |
|  | AI-ECG composite | 69.5 (66.3–72.7) | 81.6 (81.2–81.9) | 6.2 (5.7–6.7) | 99.3 (99.3–99.4) |
| PH | AI-ECG LVSD | 43.3 (40.8–46.0) | 92.6 (92.4–92.8) | 14.9 (13.9–16.0) | 98.2 (98.1–98.3) |
|  | AI-ECG LVDD | 77.1 (74.9–79.4) | 85.1 (84.8–85.4) | 13.4 (12.7–14.2) | 99.2 (99.1–99.3) |
|  | AI-ECG composite | 79.7 (77.7–81.9) | 82.5 (82.2–82.9) | 12.0 (11.4–12.7) | 99.3 (99.2–99.4) |
| CUIMC |  |  |  |  |  |
| Reduced LVEF | AI-ECG LVSD | 72.3 (71.3–73.3) | 88.9 (88.6–89.3) | 62.9 (61.9–63.9) | 92.5 (92.2–92.8) |
|  | AI-ECG LVDD | 78.4 (77.5–79.3) | 60.6 (60.0–61.1) | 34.0 (33.3–34.7) | 91.5 (91.1–91.9) |
|  | AI-ECG composite | 84.9 (84.1–85.7) | 59.0 (58.4–59.6) | 35.0 (34.3–35.6) | 93.8 (93.4–94.1) |
| VHD | AI-ECG LVSD | 51.0 (49.7–52.3) | 81.5 (81.1–81.9) | 34.1 (33.1–35.2) | 89.8 (89.4–90.2) |
|  | AI-ECG LVDD | 82.4 (81.4–83.3) | 59.2 (58.6–59.7) | 27.6 (26.9–28.2) | 94.7 (94.4–95.0) |
|  | AI-ECG composite | 84.6 (83.8–85.6) | 56.5 (56.0–57.1) | 26.8 (26.2–27.5) | 95.1 (94.8–95.4) |
| LVH | AI-ECG LVSD | 40.4 (39.3–41.5) | 80.2 (79.8–80.6) | 32.1 (31.2–33.0) | 85.3 (84.9–85.7) |
|  | AI-ECG LVDD | 73.8 (72.8–74.8) | 58.7 (58.1–59.2) | 29.3 (28.6–30.0) | 90.6 (90.2–91.0) |
|  | AI-ECG composite | 76.3 (75.4–77.2) | 56.1 (55.5–56.6) | 28.7 (28.1–29.4) | 91.1 (90.7–91.5) |
| PH | AI-ECG LVSD | 45.9 (44.3–47.3) | 79.7 (79.3–80.2) | 25.9 (25.0–26.8) | 90.5 (90.2–90.9) |
|  | AI-ECG LVDD | 77.9 (76.7–79.0) | 57.3 (56.7–57.8) | 22.0 (21.4–22.5) | 94.4 (94.0–94.7) |
|  | AI-ECG composite | 79.7 (78.6–80.7) | 54.6 (54.0–55.1) | 21.3 (20.7–21.9) | 94.6 (94.2–94.9) |

This table summarizes the phenotype-specific performance of the AI-ECG models for LVSD and LVDD across individual SHD phenotypes in the ISH and CUIMC cohorts.

AI-ECG, artificial intelligence–enabled electrocardiography; SHD, structural heart disease; LVSD, left ventricular systolic dysfunction; LVDD, left ventricular diastolic dysfunction; PPV, positive predictive value; NPV, negative predictive value; LVEF, left ventricular ejection fraction; VHD, valvular heart disease; LVH, left ventricular hypertrophy; PH, pulmonary hypertension; ISH, Incheon Sejong Hospital; CUIMC, Columbia University Irving Medical Center.

**Supplementary Table 6.** Subgroup analysis of AI-ECG composite performance for SHD detection

| **Subgroup** | **N** | **Sensitivity (%)** | **Specificity (%)** | **PPV (%)** | **NPV (%)** |
| --- | --- | --- | --- | --- | --- |
| Age ≥65 years | 18016 | 77.7 (76.3–79.0) | 74.4 (73.7–75.0) | 43.5 (42.3–44.7) | 92.9 (92.4–93.4) |
| Age <65 years | 28066 | 61.9 (59.9–64.1) | 96.1 (95.8–96.3) | 57.2 (55.1–59.2) | 96.7 (96.5–97.0) |
| Female | 22111 | 74.2 (72.5–76.0) | 85.3 (84.8–85.8) | 37.8 (36.4–39.3) | 96.5 (96.2–96.8) |
| Male | 23971 | 70.1 (68.5–71.6) | 91.2 (90.9–91.6) | 57.6 (56.1–59.1) | 94.7 (94.4–95.0) |
| BMI ≥30 kg/m² | 3983 | 70.2 (65.9–74.4) | 89.9 (88.8–90.9) | 49.1 (45.3–53.0) | 95.6 (94.9–96.3) |
| BMI <30 kg/m² | 41511 | 71.4 (70.1–72.7) | 88.5 (88.2–88.8) | 46.9 (45.9–48.0) | 95.6 (95.4–95.8) |
| **Past medical history** | | | | | |
| Hypertension | 13466 | 72.5 (70.9–74.2) | 80.0 (79.3–80.7) | 44.6 (43.1–46.1) | 92.9 (92.4–93.4) |
| Non-hypertension | 32616 | 71.2 (69.8–72.7) | 91.5 (91.2–91.8) | 49.3 (47.9–50.6) | 96.5 (96.3–96.7) |
| Diabetes mellitus | 3919 | 79.8 (77.0–82.3) | 75.1 (73.5–76.6) | 52.7 (50.2–55.3) | 91.4 (90.3–92.5) |
| Non-Diabetes mellitus | 42163 | 70.1 (68.9–71.3) | 89.4 (89.0–89.7) | 46.1 (45.1–47.2) | 95.8 (95.6–96.1) |
| Prior acute myocardial infarction | 1483 | 77.2 (73.5–80.5) | 78.9 (76.3–81.5) | 71.6 (68.0–75.1) | 83.4 (80.8–85.8) |
| Non- prior acute myocardial infarction | 44599 | 71.2 (70.1–72.4) | 88.5 (88.2–88.9) | 45.3 (44.2–46.4) | 95.8 (95.7–96.0) |
| Prior heart failure | 8306 | 80.9 (79.6–82.3) | 67.3 (65.9–68.6) | 60.6 (59.1–62.1) | 85.0 (83.9–86.1) |
| Non-prior heart failure | 37776 | 60.9 (59.2–62.7) | 91.4 (91.1–91.7) | 35.0 (33.7–36.4) | 96.9 (96.7–97.0) |
| Atrial fibrillation | 4855 | 81.1 (79.3–83.0) | 61.1 (59.4–62.8) | 52.0 (50.1–53.9) | 86.2 (84.7–87.5) |
| Non-atrial fibrillation | 41227 | 68.1 (66.6–69.5) | 90.7 (90.4–91.0) | 45.2 (44.0–46.5) | 96.2 (96.0–96.4) |
| Chronic kidney disease | 1526 | 90.2 (88.0–92.5) | 50.8 (47.2–54.1) | 61.0 (57.9–63.9) | 85.9 (83.0–89.0) |
| Non-chronic kidney disease | 44556 | 69.3 (68.1–70.6) | 89.1 (88.8–89.4) | 45.4 (44.3–46.4) | 95.7 (95.5–95.9) |
| Stroke | 3801 | 78.3 (75.1–81.5) | 75.4 (74.0–76.9) | 40.5 (37.8–43.2) | 94.2 (93.3–95.1) |
| Non-stroke | 42281 | 71.0 (69.7–72.2) | 89.4 (89.1–89.7) | 48.4 (47.2–49.5) | 95.7 (95.5–95.9) |
| **ECG rhythm** |  |  |  |  |  |
| Sinus rhythm | 41931 | 66.8 (65.4–68.3) | 90.6 (90.3–90.9) | 43.6 (42.4–44.9) | 96.2 (96.0–96.4) |
| Atrial fibrillation | 3331 | 82.5 (80.5–84.3) | 51.9 (49.8–54.2) | 56.8 (54.7–59.0) | 79.4 (77.2–81.6) |
| Non-atrial fibrillation | 42751 | 68.3 (67.0–69.6) | 90.1 (89.8–90.4) | 44.3 (43.1–45.5) | 96.1 (95.9–96.3) |
| Left bundle branch block | 586 | 94.3 (91.6–96.7) | 37.8 (32.0–43.5) | 63.5 (59.3–67.8) | 85.2 (78.8–91.0) |
| Non-left bundle branch block | 45496 | 70.6 (69.4–71.7) | 88.7 (88.4–89.0) | 46.3 (45.3–47.4) | 95.6 (95.4–95.8) |
| Right bundle branch block | 2122 | 78.2 (74.6–81.8) | 76.5 (74.4–78.7) | 50.7 (47.0–54.3) | 91.9 (90.5–93.3) |
| Non-right bundle branch block | 45496 | 71.2 (69.9–72.3) | 88.8 (88.5–89.2) | 46.9 (45.7–48.0) | 95.7 (95.5–95.9) |
| Left ventricular hypertrophy | 10079 | 77.8 (76.1–79.6) | 79.4 (78.5–80.3) | 50.7 (49.0–52.4) | 92.9 (92.4–93.5) |
| Non-left ventricular hypertrophy | 36003 | 68.2 (66.7–69.7) | 90.5 (90.2–90.8) | 45.1 (43.8–46.3) | 96.2 (95.9–96.4) |
| Pacemaker rhythm | 312 | 90.6 (86.0–94.8) | 28.3 (20.9–35.7) | 57.2 (51.2–63.5) | 74.0 (62.0–84.7) |
| Non-pacemaker rhythm | 45770 | 71.3 (70.1–72.5) | 88.6 (88.3–88.9) | 46.9 (45.9–48.0) | 95.6 (95.4–95.8) |

AI-ECG, artificial intelligence–enabled electrocardiography; SHD, structural heart disease; PPV, positive predictive value; NPV, negative predictive value; BMI, body mass index.
